# Epidemiology and Management of Malignancies in Patients with Inborn Errors of Immunity - An ESID Registry Study of 19,959 Patients

**DOI:** 10.1101/2025.08.26.25334420

**Authors:** Delfien J.A. Bogaert, Christina H. Wolfsberger, Andishe Attarbaschi, Jonathan Gathmann, Klaus Warnatz, Gabriele Mueller, Anna Mukhina, Stephan Rusch, ESID Registry Working Party, Gerhard Kindle, Joris M. van Montfrans, Markus G. Seidel

**Affiliations:** Division of Pediatric Hematology-Oncology, Department of Pediatrics, University Hospital Brussels (UZ Brussel), Brussels, Belgium; Styrian Children’s Cancer Research Unit for Cancer and Inborn Errors of the Blood and Immunity in Children, Graz, Austria; Division of Neonatology, Department of Pediatrics and Adolescent Medicine, Medical University of Graz, Graz, Austria; St. Anna Children’s Hospital, Medical University of Vienna, Vienna Austria; St. Anna Children’s Cancer Research Institute, Vienna, Austria; Institute for Immunodeficiency, Center for Chronic Immunodeficiency, Medical Center, University of Freiburg, Faculty of Medicine, University of Freiburg, Freiburg, Germany; Centre for Biobanking FREEZE, Medical Center-University of Freiburg, Faculty of Medicine, University of Freiburg, Freiburg, Germany; Clinic of Rheumatology and Clinical Immunology, Center for Chronic Immunodeficiency (CCI), Medical Center, Faculty of Medicine, Albert-Ludwigs-University of Freiburg, Germany; Dmitry Rogachev National Medical Center of Pediatric Hematology, Oncology and Immunology, Ministry of Healthcare of the Russian Federation, Moscow, Russian Federation; Treuhandstelle, Trusted Third Party, Faculty of Medicine, Medical Center-University of Freiburg, Freiburg, Germany; Department of Pediatric Immunology and Infectious Diseases, Wilhelmina Children’s Hospital, University Medical Centre, Utrecht, The Netherlands; Division of Pediatric Hematology-Oncology, Department of Pediatrics and Adolescent Medicine, Medical University of Graz, Graz, Austria

**Keywords:** inborn errors of immunity (IEI), primary immunodeficiency (PID), primary immune disorder (PID), malignancy, cancer predisposition syndrome (CPS), ESID registry

## Abstract

**Background:** Inborn errors of immunity (IEI), or primary immune disorders (PID), predispose individuals to infections, autoimmunity, inflammation, allergy, and malignancy. Malignancies are a major cause of morbidity and mortality in IEI/PID patients, with poorer outcomes compared to the general population.

**Objectives:** This European Society for Immunodeficiencies Registry (ESID-R) study aimed to determine the frequency and types of malignancies in IEI/PID patients and to assess clinical management approaches across Europe.

**Methods:** Descriptive analyses were performed on malignancy data within each IEI category. Additionally, an ESID-R survey (05/2022–03/2024) collected data on management strategies and challenges.

**Results:** Of 19,959 IEI/PID patients, 1,783 (8.9%) developed malignancies, of whom 27.1% presented malignancy as first manifestation of IEI/PID. A total of 1,210 malignancies were specified; B-cell non-Hodgkin lymphoma was most common (24.2%). Detailed malignancy–IEI/PID association maps are provided. Predominantly antibody deficiencies accounted for 59.1% of malignancy cases, with a higher median age at first malignancy (43.6 years) compared to other categories, *e.g*., combined immunodeficiencies with syndromic or associated features (11.7 years). Survey findings revealed oncological treatment was modified due to IEI/PID in 21.5% of cases, with assumed negative impacts of IEI/PID on complications and outcomes (in 27.4% and 30.7%, respectively). IEI/PID influenced transplant decisions in 16.5% of cases. Management practices like interdisciplinary decision finding and guideline availability were recorded.

**Conclusion:** This study provides comprehensive epidemiological data on malignancies in IEI/PID, highlighting the need for tailored screening and management. Survey results emphasize the real-world challenges and support the development of IEI/PID-specific oncologic surveillance guidelines and treatment strategies.

**Clinical Implications:** Overall, ca. 9% of IEI/PID patients develop malignancies —often as first presentation and highly varying between IEI/PID categories— most commonly B-cell non-Hodgkin lymphoma. High oncological treatment modification rates highlight the need for IEI/PID-specific cancer surveillance and interdisciplinary management strategies.

**Capsule Summary:** This large ESID-R study quantifies malignancy burden in IEI/PID, identifies disease-specific patterns, and reveals management modifications and challenges, underscoring the urgent need for dedicated oncologic surveillance guidelines and tailored treatment strategies in this vulnerable population.

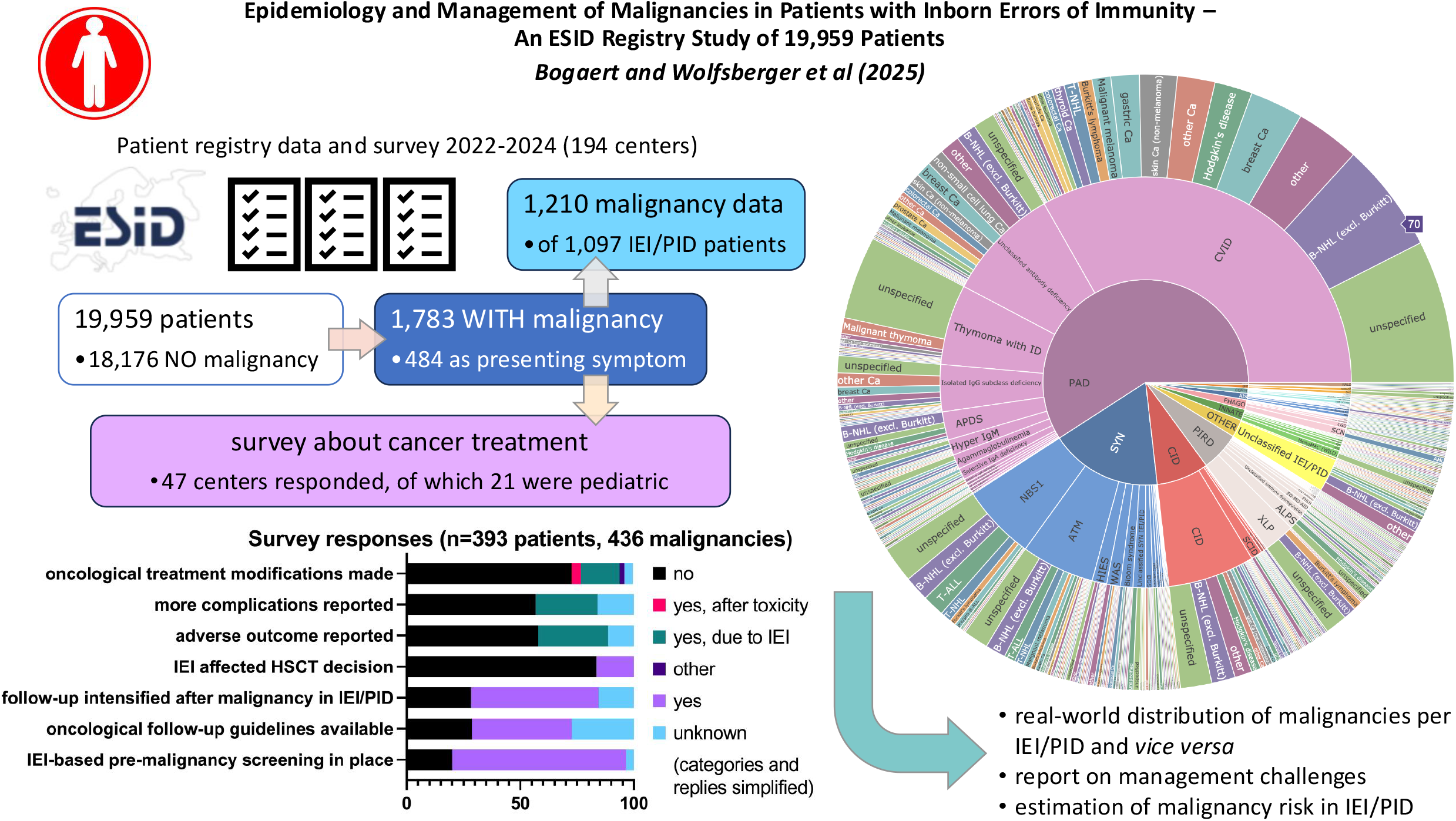

## INTRODUCTION

Inborn errors of immunity (IEI), or primary immune disorders (PID), comprise a heterogeneous group of disorders characterized by varying defects in the immune system and are currently associated with mutations in 508 different genes, as well as 17 phenocopies ^1^. Individual IEI/PID subtypes are usually rare or extremely rare, but, collectively, they represent a relevant global health burden ^2 3^. Besides an increased susceptibility to infections IEI/PID may present with autoimmunity, autoinflammation, allergy and malignancies; they are increasingly recognized as cancer predisposition syndromes ^4 5^. Multiple mechanisms are involved in the development of malignancies in IEI/PID patients. Next to the historical concept of impaired immunosurveillance as an important cancer driver, recent work has demonstrated that the same molecular defect causing the IEI/PID can also directly promote oncogenesis ^6 7 8^. These factors intrinsic to the IEI/PID encompass, for example, defects in hematopoietic cell differentiation, lymphocyte signaling, or DNA repair. In addition, extrinsic factors indirectly linked to the IEI/PID can contribute to carcinogenesis, such as chronic inflammation and inadequately controlled infection with transforming pathogens ^6 7 8^.

The overall relative cancer risk in individuals with IEI/PID is estimated to be 1.42 to 2.3-fold higher than in the general population ^9 10 11^. However, considerable variation exists between IEI/PID subtypes in the extent, type and at which age they develop malignancies ^1 12^. Lymphomas—primarily mature B-cell non!zlHodgkin lymphomas (B-NHL)—are the most common malignancy in IEI/PID. Solid tumors and leukemias also occur but are considerably less frequent. Specifically, B-NHL accounts for approximately 48–50% of cancers in IEI cohorts, with diffuse large B-cell lymphoma representing a significant share^13^. Importantly, malignancy may be the first clinical manifestation of an underlying IEI/PID, especially when the disorder has a mild, atypical or late-onset clinical presentation or its treatment is associated with unusual toxicities ^14 5^. Generally, oncological outcomes in IEI/PID patients are considered to be worse than in immunocompetent patients, and the occurrence of a malignancy in patients with IEI/PID substantially diminishes survival probabilities as compared to those without a malignancy. This is mainly attributed to increased treatment-related toxicities, reduced efficacy of standard oncological regimens, and/or higher risks of relapse and secondary malignancies ^12 15 16^. Despite growing awareness of the elevated cancer risk in IEI/PID patients and associated poorer outcomes, management remains highly variable ^12 15 16 17 18 19 20^. Currently, cancer is the second leading cause of death in IEI/PID patients, following infections ^12^. Two key strategies may help improve this unfavorable situation: first, by standardizing surveillance screening and preventive care; and second, by developing up-front adapted treatment protocols. To support these strategies with robust evidence, especially considering the broad clinical spectrum and rarity of many IEI/PID subtypes, additional data collected on a large-scale is required. In this study, we retrospectively analyzed the types, management, and outcomes of malignancies in IEI/PID patients using data from the European Society for Immunodeficiencies (ESID) registry, supplemented by a complementary survey conducted among ESID registry documenting centers. With this dual approach, we aimed to provide a more comprehensive picture of cancer in IEI/PID, addressing gaps in epidemiological knowledge and real-world clinical management challenges.

## METHODS

### Study design

Data on malignancies in IEI/PID patients were retrospectively retrieved from the ESID registry (Level 1, L1) and through an additional survey (Level 2, L2). The ESID registry is an international database on IEI/PID patients collecting their clinical and laboratory features, genetic findings, and treatments ^3^. L1 of the ESID registry gathers a standardized core set of data, including demographic information, clinical and genetic diagnoses, and whether a malignancy occurred during the patient’s lifetime. Registrants are requested to update level one data annually. All registered patients provided written informed consent themselves or by their parents or legal representatives at their treating centers, in accordance with the ESID agreement and their local ethics committees (based on latest amendment approval from IRB 00002556/24-334ex11/12) ^21 22^. Levels 2 and 3 of the ESID registry allow the implementation of research projects that collect additional information to address specific questions in either a selected group of patients/disorders or a single IEI/PID. For this study, additional L2 data were obtained from registered patients who had developed cancer. By means of an electronic survey, details on malignancy type, treatment, outcome and follow-up were collected. All ESID registry documenting centers were invited to participate in this L2 survey. A call for participation was emailed directly to these centers (*n*=194), posted on the ESID website, and announced at relevant conferences. Furthermore, a notification was displayed within the online ESID registry interface. Respondents were asked to complete the survey in consultation with the treating oncologists. The survey was done with REDCap (Research Electronic Data Capture), hosted at University Medical Centre Freiburg ^23 24^. The L2 survey was open from May 1, 2022, to March 31, 2024. Survey questions are provided in the Online Repository.

### ESID registry L1 study population

In L1 of the ESID registry, the malignancy status is recorded in either a field for presenting manifestations or in the current status report, which indicates whether the patient has developed a malignancy in his/her/its lifetime - and if so, it asks to specify the type. The age at diagnosis of the malignancy is not documented in L1. In the current study, a subgroup of these patients of whom the malignancy type was sufficiently documented at L1 (1,097/1783 (61.5%) patients with 1,210 malignancies) was further analyzed for the association between IEI/PID subtype/category and type of malignancy (Figure 1A). IEI/PID subtypes were categorized based on the latest classification of the International Union of Immunological Societies (IUIS): **I**., immunodeficiencies affecting cellular and humoral immunity or combined immunodeficiencies (CID), **II**., combined immunodeficiencies with associated or syndromic features, **III**., predominantly antibody deficiencies (PAD), **IV**., diseases of immune dysregulation (sometimes referred to as primary immune regulatory disorders, PIRD; although including more entities than those, *e.g*., also hemophagocytic lymphohistiocytosis disorders), **V**., congenital defects of phagocyte number or function, **VI**., defects in intrinsic and innate immunity, **VII**., autoinflammatory disorders, VIII., complement deficiencies, **IX**., bone marrow failure disorders, **X**., phenocopies of IEI/PID associated with autoantibodies or somatic variants, and unspecified IEI/PID ^1^.

**Figure 1.**
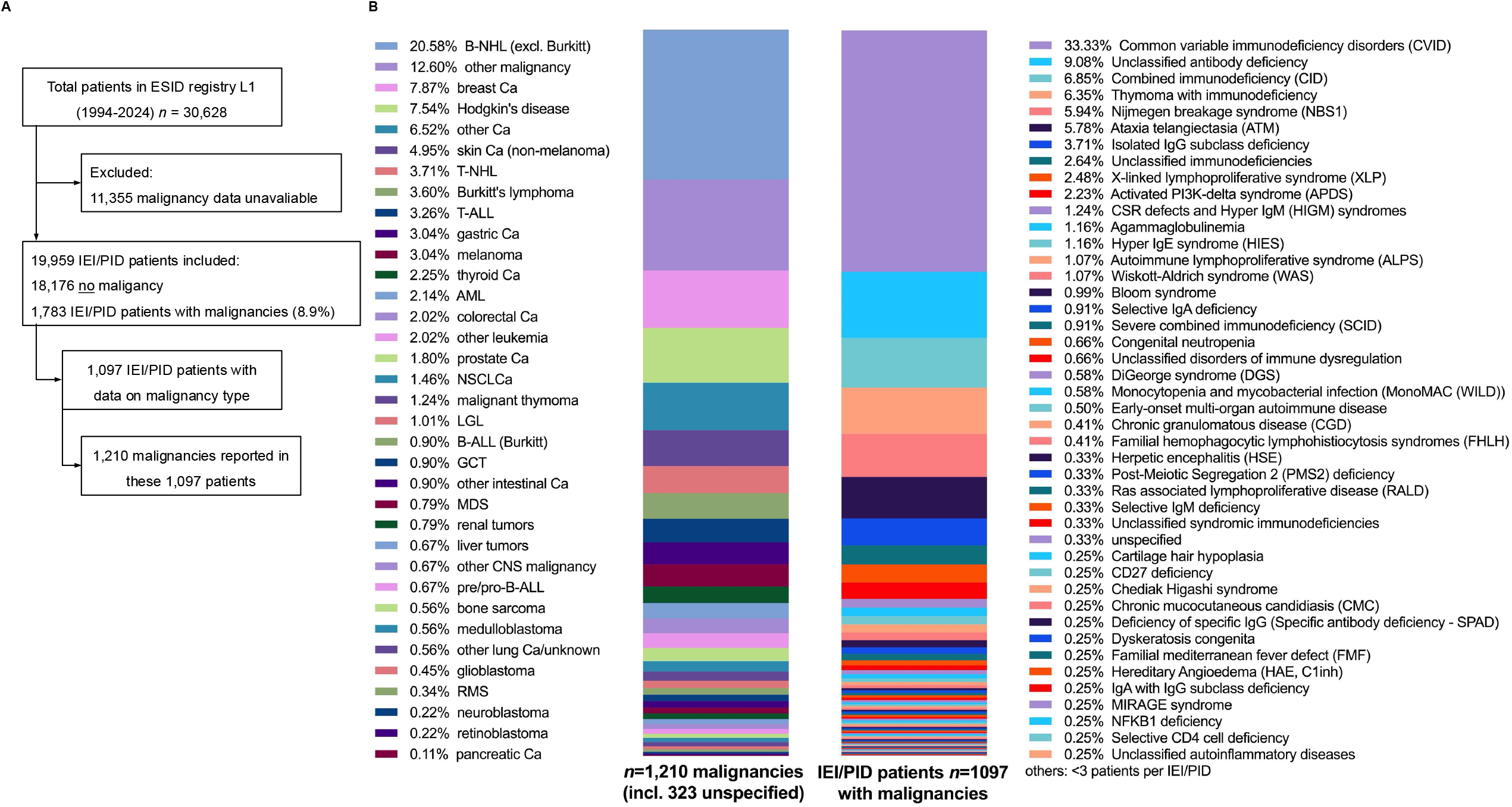
ESID registry L1 cohort. **A**, Flowchart showing the selection of the ESID registry L1 cohort on IEI/PID patients with malignancies. **B**, Distribution of malignancy types in IEI/PID patients (left) and distribution of IEI/PID subtypes in patients with malignancies (right) in the analyzed ESID registry L1 cohort. The link between malignancy subtypes and IEI/PID categories and diagnoses together with absolute numbers per diagnosis are graphically shown in an interactive hierarchical pie chart with mouse-over and click-to-zoom function at https://esid.org/html-pages/Suppl_figure_ESID_sunburst_malignancy_17.html) (IEI/PID-centered view) and at https://esid.org/html-pages/Suppl_figure_ESID_sunburst_malignancy_rev_29.html (malignancy-centered view), while the figures E1 and E2 in the Supplementary material show only screenshots of the html files. Ca, carcinoma; IEI, inborn errors of immunity; L1, level 1; PID, primary immunodeficiency.

### Statistical analysis

Statistical analysis was done using GraphPad Prism (version 10.4.2) for L1 registry data, and IBM SPSS Statistics (version 26.0) for L2 survey data. The interactive sunburst charts were designed with Plotly Open Source Graphing Library for Python (https://plotly.com/python/). In the L2 survey analysis, missing data (non-responses) were excluded. The answer option ‘unknown’ was considered a valid response and included in the statistical analysis.

## RESULTS

### Malignancies reported in the ESID registry L1 study population

As recently published in the ESID registry 1994-2024 report, information on the malignancy status was available for 19,959 of 30,628 registered IEI/PID patients, where 1,783 out of 19,959 patients (8.9%) had a history of malignancy; 484 (27.1%) of whom as first presenting symptom ^3^. The present study examined the association between IEI/PID categories and subtypes and the types of malignancy in 1,097 IEI/PID patients with a total of 1,210 malignancies recorded in the ESID registry L1 (Figure 1). Interactive html Figures E1 and E2 in the Online Repository show hierarchical association pie charts with zoom-in functionality and provide more detail regarding proportions and absolute numbers of which malignancy types occurred in which IEI/PID (Figure E1: https://esid.org/html-pages/Suppl_figure_ESID_sunburst_malignancy_17.html) and *vice versa* (Figure E2: https://esid.org/html-pages/Suppl_figure_ESID_sunburst_malignancy_rev_29.html). Overall, lymphomas were the most frequently reported malignancies, accounting for 36.3 % of cases. Among these, B-NHL (excluding Burkitt lymphoma) represented 20.6%, Hodgkin’s disease 7.5%, T-NHL and Burkitt lymphoma 3.7% and 3.6%, respectively (Figure 1). Solid tumors were also highly prevalent, with skin cancers (both melanoma and non-melanoma, 8.0%) and breast cancer (7.9%) being the most commonly reported types. Leukemias accounted for only 8.0% of malignancies in the ESID registry cohort (Figure 1B, Figure E2). Notably, certain IEI/PID subtypes were associated with specific malignancy types. While B-NHL (excluding Burkitt lymphoma) was the most prevalent malignancy type across many IEI/PID subtypes, myeloid neoplasms were more frequently observed in phagocyte disorders, DNA repair disorders, and bone marrow failure syndromes (Figure E2). PAD, and more specifically common variable immunodeficiency (CVID), were the most frequently recorded IEI/PID category and subtype, respectively, associated with malignancy (Figures E1 and E2). PAD was reported in 59.1% of all cancer patients in the ESID registry, with CVID accounting for 33.3% (Figure 1B).

### Estimation of malignancy risk of selected IEI/PID diagnoses

Replies to the registry level 1 question “*Did the patient ever suffer from malignancy?*” allow an estimation of the overall risk distribution of malignancies across various IEI/PID. Table 1 shows the minimum and estimated prevalence of current or past malignancy across selected IEI/PID subtypes, including CVID (malignancies reported in 404 patients out of 6686; unknown/not reported in 35%; prevalence 6.0-9.3%), unclassified PAD (UnPAD; 110/2474; unknown 35%; prevalence 4.4-6.9%), Ataxia telangiectasia (AT; 70/792; unknown 34%; risk 8.8-13.4%), CID not further specified (CID n.f.s.; 83/789; unknown 33%; prevalence 10.5-15.7%), autoimmune lymphoproliferative syndrome (ALPS; 13/428; unknown 20%; prevalence 3-3.8%), Nijmegen breakage syndrome (NBS; 72/256; unknown 34%; prevalence 28.1-42.6%), and activated PI3K delta syndrome (APDS; 27/173; unknown 35%; prevalence 15.6-24.1%).

**Table 1.** Estimated prevalence of current or history of malignancy in selected IEI/PID.

|  | **CVID** | **UnPAD** | **A.T.** | **CID n.f.s.** | **ALPS** | **NBS** | **APDS** |
| --- | --- | --- | --- | --- | --- | --- | --- |
| *Total in ESID registry<sup>1</sup> n=* | 6686 | 2474 | 792 | 789 | 428 | 256 | 173 |
| *Malignancy reported n=* | 404 | 110 | 70 | 83 | 13 | 72 | 27 |
| *Estimated reporting fraction<sup>2</sup>* | 0.65 | 0.65 | 0.66 | 0.67 | 0.80 | 0.66 | 0.65 |
| *Minimum prevalence (%)* | 6.0 | 4.4 | 8.8 | 10.5 | 3.0 | 28.1 | 15.6 |
| *Estimated prevalence (%)* | 9.3 | 6.9 | 13.4 | 15.7 | 3.8 | 42.6 | 24.1 |

<sup>1</sup>, data from ESID registry 2025 report on 30,628 patients; selected IEI/PID diagnoses listed in descending order, according to *Kindle and Alligon et al. 2025[3]*
<sup>2</sup>, proportion of patients of whom the question “Did the patient ever suffer from malignancy?” was answered (“Yes” or “No”) in the corresponding IUIS category in the ESID level 1 registry, according to *Kindle and Alligon et al. 2025[3]*

### Characteristics of the L2 survey cohort

Overall, colleagues from 47 ESID registry documenting centers responded to the call for the L2 survey study (24.2% response rate). Based on the names of the documenting centers, 21 were exclusively pediatric and 3 exclusively adult. For the remaining 23 centers, it was unclear whether they served pediatric patients, adults, or both. The survey was successfully completed for 393 patients, *i.e*., 35.8% (393/1,097) of the registered IEI/PID patients with cancer of whom data on malignancy type was available (Figure 2A). The demographics of the survey cohort are shown in Table 2. Comparable with the IEI/PID distribution of the L1 cohort of patients with malignancies –and thus representative–, the majority of patients in the L2 survey cohort had PAD as underlying condition, followed by combined immunodeficiencies with associated or syndromic features, and diseases of immune dysregulation (50.1%, 24.3%, and 9.5%, respectively). Of note, no patients with bone marrow failure syndromes were registered in the L2 survey. The median duration of follow-up was 11.5 years (range 0-54 years). A total of 436 malignancies were reported in the survey cohort, with 39 patients who had developed more than one malignancy. The median age at diagnosis of the first malignancy was 29.0 years (range 0-86 years), and of the second or higher malignancy 54.0 years, both ranging from early childhood to late adulthood (Table 2). The first malignancy was reported to be the presenting feature of the IEI/PID (malignancy diagnosed prior to or simultaneously with IEI/PID) in about 30% of the survey cohort. This was more often indicated in children (< 18 years of age) than in adults: in 38.6% of pediatric cases versus 25.6% of adult cases was the malignancy reported as presenting feature of the IEI/PID. The age at diagnosis of the first malignancy differed substantially between the categories of the underlying IEI/PID, with IEI/PID with syndromic features showing the earliest presentation (median 11.7 years, *n*=95), followed by disease of immune dysregulation (17.8 years, *n*=37), while that of PAD was 43.6years (*n*=196; Figure 2D).

**Table 2.** Demographics of the ESID registry L2 sub-study/survey cohort (n=393).

| **Gender** | **Number (percentage)** |  |
| --- | --- | --- |
| Male | 210 (53.4%) |  |
| Female | 183 (46.6%) |  |
| **IEI/PID category<sup>o</sup>** | **Number (percentage)** | **Percentage in L1 cohort (n=1783 patients)<sup>2</sup>** |
| Immunodeficiencies affecting cellular and humoral immunity | 33 (8.4%) | 6.3% |
| Combined immunodeficiencies with associated or syndromic features | 95 (24.3%) | 17.5% |
| Predominantly antibody deficiencies | 196 (50.1%) | 58% |
| Diseases of immune dysregulation | 37 (9.5%) | 8.4% |
| Congenital defects of phagocyte number or function | 2 (0.5%) | 4.2% |
| Defects in intrinsic and innate immunity | 2 (0.5%) | 1.8% |
| Autoinflammatory disorders | 6 (1.5%) | 0.7% |
| Complement deficiencies | 6 (1.5%) | 0.84% |
| Bone marrow failure disorders | 0 | 0.4% |
| Phenocopies of inborn errors of immunity | 6 (1.5%) | 0.3% |
| Unspecified IEI/PID | 8 (2.0%) | 1.6% |
| **Genetic IEI/PID diagnosis** | **Number (percentage)** |  |
| Yes | 188 (47.8%) |  |
| No | 205 (52.2%) |  |
| **First malignancy** | **Median (range) / Number (percentage)** |  |
| Age (in years) at diagnosis of first malignancy | 29.0 years (0-86 years) |  |
| First malignancy diagnosed in childhood (< 18 years of age) | 157 (39.9%) |  |
| First malignancy diagnosed in adulthood (≥ 18 years of age) | 236 (60.1%) |  |
| **Second or higher malignancy** | **Median (range)** |  |
| Age (in years) at diagnosis of second or higher malignancy | 54.0 years (5-78 years) |  |
| **Malignancy as presenting feature of IEI/PID** | **Number (percentage)** |  |
| Yes | 120 (30.8%) |  |
| No | 269 (69.2%) |  |
| **Primary malignancies per patient** | **Number (percentage)** |  |
| One | 354 (90.1%) |  |
| Two | 35 (8.9%) |  |
| Three | 3 (0.8%) |  |
| More than three | 1 (0.3%) |  |
| **Overall outcome at last follow-up** | **Number (percentage)** |  |
| Alive | 285 (72.5%) |  |
| Dead | 99 (25.2%) |  |
| Unknown | 9 (2.3%) |  |
| **Follow-up** | **Median (range)** |  |
| Duration (in years) of follow-up | 11.5 years (0-54 years) |  |
| Age (in years) at last follow-up | 37.0 years (3-87 years) |  |

<sup>o</sup>According to the IUIS classification *Poli et al. 2025* [1]<sup>1</sup>. IEI, inborn errors of immunity; L2, level 2; PID, primary immune disorder.
<sup>2</sup>, proportion of patients of whom the question “Did the patient ever suffer from malignancy?” was answered “Yes” between 10 IUIS IEI category out of all patients with malignancies reported in the ESID level 1 registry, according to *Kindle and Alligon et al. 2025* [3]

**Figure 2.**
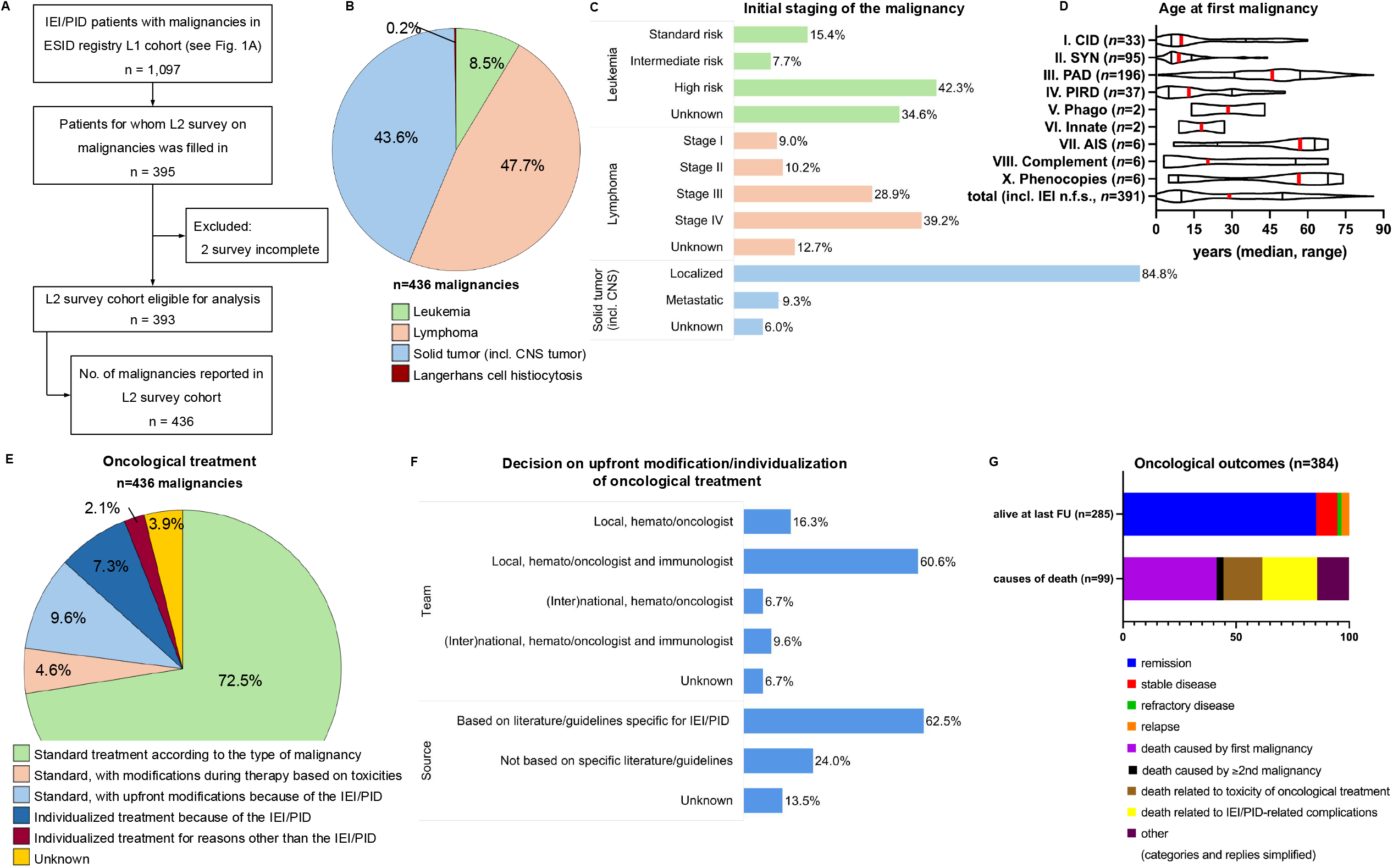
L2 survey cohort. **A**, Flowchart showing the selection of the L2 survey cohort. **B**, Overview of malignancies reported in the survey cohort. **C**, Risk groups or disease stages of the malignancies at diagnosis reported in the survey cohort. **D**, Age at diagnosis of the first malignancy across the IEI/PID categories (red marker: median, black marker: interquartile range, violins: range), the IUIS categories of IEI are shown with roman numericals and abbreviated, with “PIRD” used for all diseases of immune dysregulation, not only primary immune regulatory disorders in the strict sense. **E**, Oncological treatment and reported modification decisions. **F**, Basis for the decisions (persons, sources) on upfront treatment modifications. **G**, Oncological outcomes reported in 384 patients of the L2 survey cohort. CNS; central nervous system; IEI, inborn error of immunity; L1, level 1; L2, level 2; PID, primary immune disorder.

### Reported malignancies in the survey cohort

Of the 436 malignancies in the survey cohort, lymphomas (47.7%) and solid tumors (including brain tumors; 43.6%) were most frequently reported. Leukemias only accounted for 8.5%. One patient had multisystemic Langerhans cell histiocytosis (Figure 2B, Figure E2). The risk groups (in case of leukemias) or stages (in case of lymphomas and solid tumors) of the malignancies at diagnosis are summarized in Figure 2C and Figure E2. The distribution of malignancies across the IEI/PID categories is shown in Figure 2D and Table E1.

### Reported extrinsic cancer risk factors in the survey cohort

The survey explored which factors non-intrinsic to the IEI/PID may have contributed to the overall cancer risk. The results are summarized in the Supplementary Results and Figure E3 in the Online Repository. In summary, in up to 30.7% of malignancies, survey respondents considered extrinsic factors indirectly related to the IEI/PID to have contributed to the malignancy risk (*e.g*., chronic inflammation, oncogenic pathogen).

### Reported oncological treatment in the survey cohort

The survey respondents indicated that in the majority of malignancies (77.1%) standard oncological practice was followed, *i.e*., standard treatment with or without adaptations based on toxicities encountered during therapy (Figure 2E). However, in 16.9% of malignancies, oncological treatment was reported to be modified or completely individualized upfront because of concerns related to the underlying IEI/PID (Figure 2E). For those malignancies in which treatment was adapted upfront, it was reported that in 70.2% of cases immunologists were involved in the decision (Figure 2F, top) and that specific literature or guidelines were available in 62.5% of cases (Figure 2F, bottom). Once cancer treatment was ongoing, the involvement of immunologists was reported to be 46.1%. Allogeneic HSCT was reported to be performed after or as part of the oncological treatment in 13.3% of malignancies. The IEI/PID was considered to favor the decision of HSCT in 15.1% of patients with malignancies, while it was considered an argument against HSCT in 1.4%. Survey results concerning radiotherapy and immune therapy, and antimicrobial prophylaxis and immunological therapy during oncological treatment are provided in the Online Repository.

### Reported outcome in the survey cohort

At last follow-up, 72.5% of the survey patients were reported to be alive whereas 25.2% had deceased (2.8% unknown; Table 2). The oncological status in patients alive at last follow-up (*n*=285) is shown in Figure 2G, over 85% were reported to be in remission. Of deceased patients (*n*=99), the median age at death was 23 years (range 3-85 years). 61.6% of deaths were indicated to be due to oncological reasons: 41.4% died of a first malignancy, 3.0% of a second or higher malignancy, and 17.2% of oncological treatment toxicities (Figure 2G).

### Reported impact of the IEI/PID on oncological outcome in the survey cohort

Respondents judged that the presence of an underlying IEI/PID resulted in prolongation of the oncological treatment in 22.5% of malignancies as compared to oncological patients without IEI/PID, and negatively impacted the oncological outcome in 30.7% of malignancies (Figure 3A). Most often, the latter was reported to be due to infections or immune dysregulation related to the IEI/PID. Other reported reasons were related to pretreatment and comorbidity in the IEI/PID patient, or that the maximum oncological treatment could not be given (Figure 3B). Furthermore, in 27.4% of malignancies, complications encountered during oncological treatment were judged to be more severe compared to those in non-IEI/PID patients with similar malignancies (Figure 3C), with details of reported complications shown in Figure 3D.

**Figure 3.**
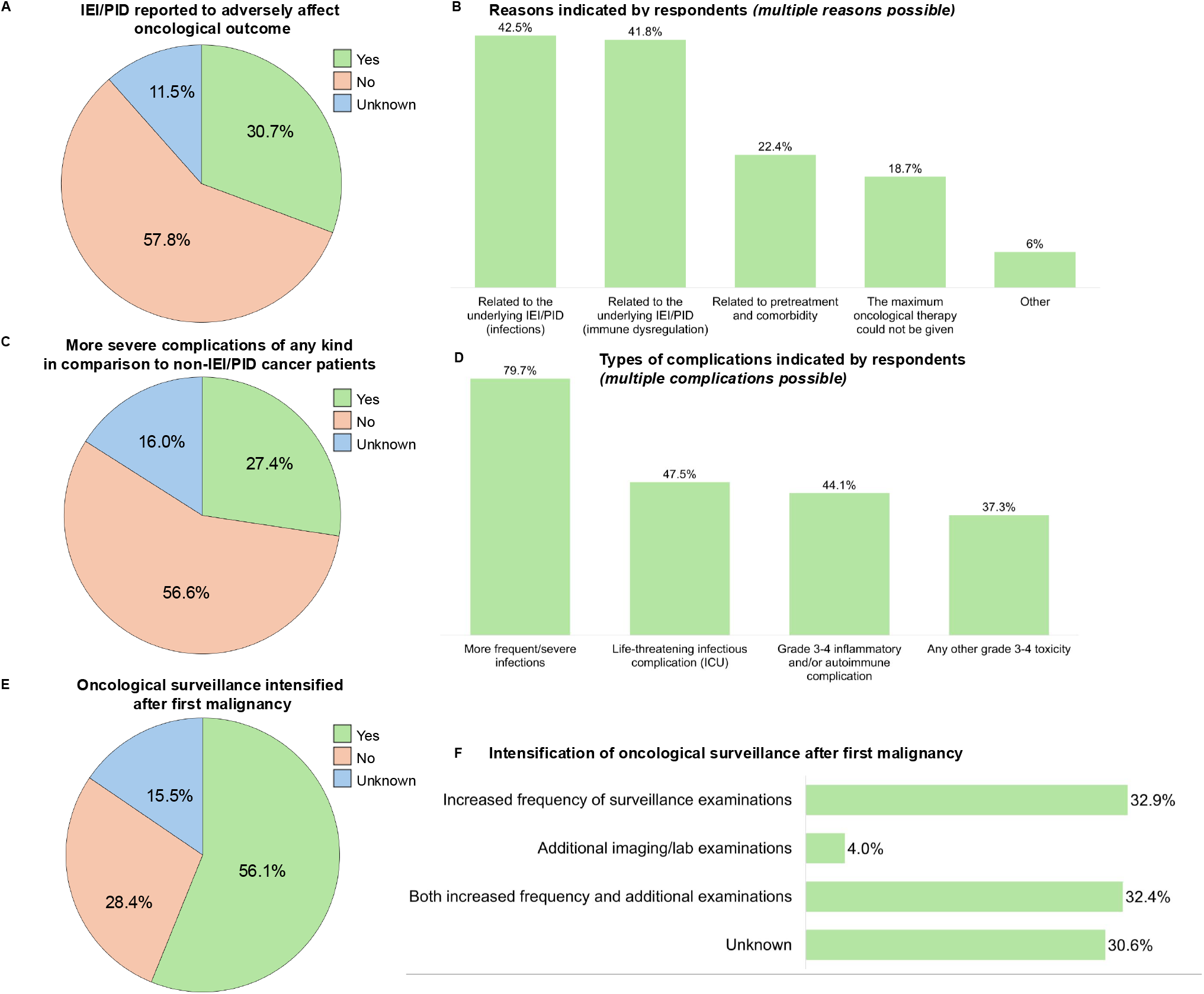
Reported effect of the IEI/PID on the oncological outcome in the survey cohort and surveillance strategies. **A**, The proportion of malignancies in which the IEI/PID was reported to adversely effect the oncological outcome. **B**, Reported reasons why the IEI/PID was judged to adversely affect the oncological outcome. **C**, The proportion of malignancies in which the IEI/PID was reported to be associated with more severe complications during oncological treatment. **D**, Types of complications reported by the survey respondents. Other grade 3-4 toxicities included anaphylactic reactions, mucositis, bone marrow toxicity (pancytopenia), gastro-intestinal toxicity, liver toxicity, lung complications, metabolic complications, neurological toxicity and multiorgan toxicity. **E**, Intensification of oncological surveillance after a first malignancy. **F**, Types of applied intensification of oncological sureveillance after a first malignancy. ICU, intensive care unit; IEI, inborn errors of immunity; PID, primary immunodeficiency.

### Reported follow-up and surveillance in the survey cohort

After successful treatment of the malignancy (n=383), follow-up was reported to be mostly done at the hematological/oncological clinic alone (44.9%) or at a combination of both the hematological-/oncological and immunological clinic (43.3%), but rarely at the immunologist alone (5.2%; unknown in 6.5%). In 56.1% of patients, oncological surveillance was reported to be intensified compared to non-IEI/PID patients after a first primary malignancy had occurred, mostly based on an increased frequency of visits with or without additional lab or imaging examinations (Figure 3E, F). Additionally, in five patients, oncological screening was reported to be intensified only after a second (*n*=4) or third (*n*=1) malignancy had occurred, also by increased frequency and/or increased number of surveillance examinations. IEI/PID-specific oncological surveillance guidelines were reported to be available in the post-malignancy setting in 43.8% of malignancies. However, in more than half of cases such guidelines were lacking or unknown to exist (Figure E4A). In contrast, IEI/PID-specific oncological surveillance guidelines were available in 76.3% of cases in the setting in which a malignancy had not yet occurred (Figure E4B).

## DISCUSSION

This retrospective study, combining data from the ESID registry (L1) with an accompanying survey (L2), constitutes one of the most extensive and detailed analyses to date on the prevalence, clinical features, and management of malignancies in patients with IEI/PID. The ESID registry offers valuable epidemiological insights into the types and distribution of malignancies across various IEI/PID categories. Notably, malignancies were reported in approximately 8.9% of patients, and in 27.1% of these cases, cancer was the initial clinical manifestation of an underlying immune disorder ^3^. The present in-depth analysis with interactive hierarchical pie charts of IEI/PID categories and diagnoses together with specified malignancy subtypes of 1210 cases (in 1097 patients) and *vice versa* provide high-resolution images of the real-world distribution of malignancies in IEI/PID, constituting robust evidence as basis for future clinical studies on specific cancer entities in IEI/PID. The incidence and median age at malignancy diagnosis varied significantly depending on the specific IEI/PID subtype, with PAD being the most common category among cancer patients, as expected due to its overall numerical predominance within the registry cohort and the population, and PAD showing the latest median age at malignancy onset. Among PAD subtypes, APDS had the highest estimated cancer risk (15.6–24.1%), exceeding that of CVID (6–9.3%) and, in comparison, that of CID n.f.s. (10.5–15.7%), but lower than that of NBS1 (28.1–42.6%). The L2 survey, conducted via REDCap between May 1, 2022, and March 31, 2024, gathered detailed information from 47 participating ESID registry centers regarding malignancy types, treatment decision-making and modifications, outcomes, and follow-up modalities. The questionnaire was developed in close collaboration with oncologists to ensure clinical relevance and completeness, and the replies draw a picture of awareness but highly varying standards.

Our findings demonstrate the variably increased cancer risks associated with different IEI/PID subtypes. PAD and CVID consistently represented the largest proportion of malignancy cases in IEI/PID as reported previously ^9 10 11 13^, which is partly attributable to their higher overall prevalence (PAD: 49.4% of patients in total ESID registry), as suggested by Bayes’ theorem, despite the intrinsically higher malignancy risk seen in other, rarer, disorders such as DNA repair defects ^8 25 26 27^. Of note, these numbers might still underestimate malignancy risks in CVID, as preexisting lymphoma is still considered an exclusion criterion for making the diagnosis of CVID. Additionally, L1 ESID registry data show that patients with PAD are older on average (median 40 years, Q1-Q3: 23-61) than those with other IEI/PID subtypes ^3^, which, due to cumulative risks, likely also contributes to the higher cancer burden observed in this category. Patient data included in the L2 survey confirmed that individuals with PAD had the highest median age at diagnosis of a malignancy (43.6 years), while CID with syndromic or associated features were the youngest subgroup at their diagnosis of a malignancy (11.7 years, versus 20 years median age [Q1-Q3: 12-29] of that category in the entire ESID registry), followed by diseases of immune dysregulation and CID. These findings are consistent with the stronger intrinsic risks for oncological transformation in CID with syndromic or associated features and DNA repair defects as well as diseases of immune dysregulation and CID, mostly due to lymphocyte differentiation, maturation, apoptosis or signaling defects ^6^. While external environmental factors and chronic infections may contribute to malignancy development, our data suggest that intrinsic mechanisms—such as impaired B-cell maturation—play a dominant role in malignancy development in IEI/PID ^6^. Although chronic infection and inflammation — such as that associated with Helicobacter pylori colonization — are well-recognized extrinsic drivers of tumorigenesis through persistent immune stimulation and impaired immune surveillance, the high prevalence of B-NHL among PAD patients suggests that intrinsic mechanisms may play a relevant role also in that category ^6 28^. Malignancy in PAD may stem from impaired B-cell development, checkpoint regulation, or DNA repair, or altered signaling, rendering B cells intrinsically susceptible to transformation, further fostered by extrinsic cues ^29^. The repeated emergence of B-NHL, even in older individuals with long-standing antigenic stimulation and chronic inflammation, points toward underlying defects in B-cell development, maturation, and genomic integrity as key contributors to malignant transformation.

Overall, our study confirms that B-NHL is the predominant malignancy in IEI/PID (24.2% incl. Burkitt lymphoma), with solid tumors such as breast (7.9%) and skin cancers (8.0%) also commonly reported – which is in line with previous studies ^12 13 30 31^. <u>In contrast</u>, Additionally, we observed relatively high rates of gastric (4.5%) and thyroid cancers (2.0%) in CVID patients, which is consistent with earlier reports ^9 11 26 27^. Leukemias were relatively rare in our cohort (8.0%), reflecting their association with specific leukemia-prone IEI/PID subtypes ^31 32^. Although a non-IEI/PID control group would be needed for conclusive evidence, the malignancy risk groups/stages reported in IEI/PID patients in our L2 sub-cohort do not seem to be more severe than those in non-IEI/PID patients.

The observation that that NBS patients exhibited a particularly high prevalence of malignancies (minimum 28.1%, estimated 42.6%) is consistent with previous research indicating an extremely high incidence of malignancies, mostly NHL, in NBS patients ^33^. Similarly, AT patients show elevated malignancy prevalence (minimum 8.8%, estimated 13.4%). Although these numbers corroborate findings from the French National Registry of Primary Immune Deficiencies ^34^, they may still underestimate the real risk of persons with AT, as they may also be underreported in the ESID registry, depending on their main clinical manifestations. Likewise, the prevalence of malignancies in CVID patients reported in the ESID registry (minimum 6.0%, estimated 9.3%) is probably an underestimation. Studies including an Italian retrospective study have found higher incidences of malignancy in CVID patients (27%) ^35^ than observed in our data. This could be due to differences in study design, patient populations and their geographical malignancy risks, diagnostic criteria, or follow-up periods and follow-up institutions, with oncological care centers unlikely to report back to the ESID registry. The Italian study also noted lymphomas as the most frequent malignancy in their CVID cohort ^35^, and malignancy was reported to be among possible initial clinical presentations of CVID ^36^, both in line with our data.

Apart from information on prevalences of malignancy types in IEI/PID patients, the L2 survey provided further insight into the clinical management and real-world challenges of treating malignancies in IEI/PID. Consistent with the larger L1 ESID registry data, malignancy was reported as the presenting feature of the IEI/PID in 30.8% of the L2 survey patients, underlining the importance of considering an underlying IEI/PID in patients newly diagnosed with malignancy ^12 14 15^. Moreover, nearly 10% of the survey patients were reported to have developed more than one malignancy, in line with the percentage recently published in a systematic review by Fekrvand et al. (10.1%) ^13^. Approximately 40% of first malignancies reported in the survey were diagnosed in childhood, which is higher than in other studies ^9 10 11^. This may partly be explained by the relatively large representation of pediatric centers in our survey. We noted that the outcomes of malignancies, with >72% alive at last follow-up, were substantially higher than those of a recent study of NHL in patients with underlying conditions ^17^. This might be an overestimation of survival; the discrepancy might be due to the fact that our L2 patient cohort was more heterogenous than the NHL study, and our retrospective data collection via a survey, although corrected for “unknowns”, might select for patients who successfully returned to immunological care after their oncological treatment (see *Limitations* below). Furthermore, the low numbers of different malignancy types in our L2 cohort did not allow a statistical comparison of different cancers.

The management of malignancies in IEI/PID patients requires a careful balance between the risk of a poorer response to standard oncological treatment, increased risk of treatment-related toxicity (especially in DNA repair defects), and an elevated risk of relapse and secondary malignancies (such as the risk of recurrent lymphoma in CVID), compared to non-IEI/PID patients ^15^. Moreover, there is considerable variation in this risk balance between the different IEI/PID subtypes ^15^. With the survey, we aimed to gain a better understanding of current practices. According to the respondents, IEI/PID-related upfront modifications of oncological treatment were made in only 16.9% of malignancies.

Among these, the decision to modify oncological treatment upfront was based on relevant literature or clinical guidelines in 62.5%. In contrast to the small proportion of malignancies for which treatment was modified upfront, the underlying IEI/PID was reported to have adversely affected oncological outcome in roughly 31%, and to be associated with more severe treatment-related complications in about 27%; with further differentiation of these replies being too granular to interpret after splitting into many sub-groups on two sides (IEI/PID and malignancies). These replies suggest that existing literature and clinical guidelines may not sufficiently cover the complexities of managing malignancies in IEI/PID patients. As for oncological surveillance strategies, it was reported in the survey that guidelines were lacking or unknown to exist in more than half of cases. Indeed, current literature includes international consensus guidelines on cancer treatment and/or surveillance for only a limited number of IEI/PID subtypes, particularly those involving DNA repair and chromosomal instability disorders (e.g. Fanconi anemia, dyskeratosis congenita, AT, NBS, Bloom syndrome, CMMRD) ^37 38 39 19^. For several other IEI/PID subtypes, such as Shwachman-Diamond syndrome, GATA2 deficiency, and SAMD9/9L syndromes, general recommendations exist, but evidence is insufficient to establish international consensus on, for example, the frequency and extent of surveillance examinations ^40 41 42 43^. However, for many IEI/PID subtypes, including CVID and lymphoproliferative disorders, no formal recommendations or guidelines are available ^44 45 46^. In these cases, clinical practice relies on case reports, small series, or expert opinion. International efforts are needed to compile existing knowledge and develop at least a minimum set of recommendations for these IEI/PID subtypes. Finally, the survey indicated varying levels of collaboration between hematologists/oncologists and immunologists in the treatment and follow-up of IEI/PID patients with malignancy, suggesting that further steps are needed to strengthen multidisciplinary care.

### Strengths and limitations

The main strengths of this study are the use of the large, multicenter dataset provided by the ESID registry as well as the addition of a targeted survey to gather real-world data on malignancy management and outcomes in IEI/PID patients. However, several limitations must be acknowledged. The retrospective, registry-based design is inherently subject to reporting bias, missing data, and inconsistencies in detail, which can lead to a loss of information regarding malignancy subtypes, treatment timelines, or disease outcomes. Additionally, registry coverage may be incomplete, as some IEI/PID patients might be registered in national databases or in registries of related medical disciplines, such as the European Working Group of Myelodysplastic Syndromes and Severe Aplastic Anemia in Childhood (EWOG-MDS/SAA) or the European Society for Blood and Marrow Transplantation (EBMT) and not in the ESID registry. We noted a clear underrepresentation of patients with DNA-repair disorders and bone marrow failure syndromes in the ESID registry— conditions known to carry a high malignancy risk, particularly for myelodysplastic syndromes (MDS), leukemia, and T-cell lymphoblastic lymphoma. Furthermore, despite the relatively well-balanced distribution of IEI/PID diagnoses within the L2 malignancy survey cohort compared to the larger L1 registry cohort of IEI/PID patients with malignancies, the relatively low response rate to the survey (24.2% of all ESID registry centers) may limit the generalizability of the survey-based findings. Finally, inherent in the nature of a survey, several questions in the survey explored only the clinicians’ perceptions regarding management and outcomes without the need to substantiate their replies by facts and medical reports; as such, parts of the survey analysis reflect subjective clinical judgement and should be interpreted with this limitation in mind.

## CONCLUSION

In conclusion, the presented epidemiological data on the distribution of malignancy types across IEI/PID diagnoses, based on ESID registry L1 data, provides a valuable resource for clinicians and serves as a foundation for developing future clinical research studies and clinical management guidelines, including recommendations for IEI/PID screening at diagnosis of a malignancy, especially in B-NHL. The clinical need for IEI/PID-specific oncological treatment and surveillance protocols is underlined by the survey results, which provided real-world insights into the management challenges encountered in this complex patient population.

## Data Availability

All data produced in the present work are contained in the manuscript.

## Acknowledgements

The authors thank Mickaël Alligon, Paris, and Benedikt N. Seidel, Munich, for help with the programming of the interactive sunburst chart.

## ESID Registry Working Party (AND malignancy survey collaborators at the end of the list)

Abd Elaziz, Dalia; Abdelkader, Sohilla Lofty M.; Abitbol, Avigaelle; Abolhassani, Hassan; Abrahim, Lalash; Abuzakouk, Mohamed; Accardo, Pietro Andrea; Achir Moussouni, Nabila; Afonso, Veronica; Agyeman, Philipp; Ahlmann, Martina; Aiuti, Alessandro; Akl, Abla; Aksu, Güzide; Albers, Kim; Albert, Michael H.; Alecsandru, Meda Diana; Aleinikova, Olga; Aleshkevich, Svetlana; Alkady, Radwa Salah Eldeen Youssif; Allende, Luis; Alligon, Mickaël; Allwood, Zoe; Alsina, Manrique de Lara Laia; Ambrosch-Barsoumian, Daniela; Ameshofer, Lisa; Amour, Kenza; Anadol, Evrim; Ananthachagaran, Ariharan; Andriamanga, Chantal; Andris, Julia; Andritschke, Karin; Angelini, Federica; Ankermann, Tobias; Apel, Katrin; Arami, Siamak; Ardeniz, Ömür; Arkwright, Peter; Arnold, Karina; Ascherl, Rudolf; Assam, Najla; Assia-Batzir, Nurit; Atschekzei, Faranaz; Aumann, Sybille; Aumann, Volker; Aurivillius, Magnus; Ausserer, Bernd; Avcin, Tadej; Aydemir, Sezin; Aygören-Pürsün, Emel; AyvazDeniz, Cagdas; Azzari, Chiara; Babacar, Lo; Bach, Perrine; Bachmann, Sophie; Bader, Peter; Bakhtiar, Shahrzad; Bancé, Renate; Bangs, Catherine; Barfusz, Katerina; Baris, Safa; BarlanIsil, B; Bartsch, Michaela; Baselli, Lucia Augusta; Batlle-Maso, Laura; Baumann, Helge; Baumann, Ulrich; Baumeister, Veronika; Baxendale, Helen; Bazen, Suzanne; Beaurain, Beatrice; Beauté, Julien; Bechar-Makhloufi, Mounia; Beck, Norbert; Becker, Brigitta; Becker, Christian; Behrends, Uta; Beider, Renata; Beier, Rita; Belkacem, Amel; Belke, Luisa; Bellert, Sven; Belohradsky, Bernd H.; Ben-Bouzid, Aouatef; Benoît, Vincent; BenSlama, Lilia; Berdous-Sahed, Thamila; Bergils, Jan; Berglöf, Anna; Bergman, Peter; Bernat-Sitarz, Katarzyna; Bernatoniene, Jolanta; Bernatowska, Ewa; Bernbeck, Benedikt; Bertolini, Elena; Bethune, Claire; Beuckmann, Kai; Bhole, Malini; Biegner, Anika-Kerstin; Bielack, Stefan; Bienemann, Kirsten; Bigl, Arndt; Bigorgne, Amélie; Bijl, Marc; Binder, Nadine; Bitzenhofer-Grüber, Michaela; Blanchard Rohner, Geraldine; Blank, Dagmar; Blattmann, Claudia; Blau, Julia; Blaziene, Audra; Blazina, Stefan; Bloomfield, Markéta; Blume, Roswitha; Boardman, Barbara; Bode, Sebastian; Boelens, Jaap-Jan; Boesecke, Christoph; Bogaert, Delfien; Bogner, Johannes; Bohynikova, Nadezda; Booth, Claire; Bordon, Victoria; Borkhardt, Arndt; Börries, Melanie; Borte, Michael; Borte, Stephan; Bossaller, Lukas; Bossard, Madeleine; Botros, Jeannet; Boucherit, Soraya; Boutros, Jeannette; Boyarchuk, Oksana; Boyman, Onur; Boztug, Kaan; Branco Pereira da Silva, Sara; Braschler, Thomas; Bravo, Sophie; Bredius, Robbert; Bright, Philip; Brito de Azevedo Amaral, Carolina; Brodszki, Nicholas; Brodt, Grit; Brolund, Allan; Brosselin, Pauline; Brummel, Bastian; Brun-Schmid, Sonja; Brunner, Jürgen; Bruns, Roswitha; Buchta, Christina; Buck, Dietke; Bücker, Aileen; Buckland, Matthew; Bührlen, Martina; Burdach, Stefan; Burns, Siobhan; Burton, Janet; Caminal, Luis; Cancrini, Caterina; Candotti, Fabio; Canessa, Clementina; Cant, Andrew J; Cantoni, Nathan; Capilna, Brindusa; Caracseghi, Fabiola; Caragol, Isabel; Carbone, Javier; Carrabba, Maria; Casanova, Jean-Laurent; Chamberlain, Latanya; Chandra, Anita; Chantrain, Christophe; Chapel, Helen; Chassot, Julie; Chee, Ronnie; Chinello, Matteo; Chopra, Charu; Chovancova, Zita; Christmann, Martin; Chrzanowska, Krystyna; Ciznar, Peter; Claes, Karlien; Classen, Carl Friedrich; Classen, Martin; Cochino, Alexis-Virgil; Condliffe, Alison M.; Corbacioglu, Selim; Cordeiro, Ana Isabel; Cordier Wynar, Donatienne; Core, Claire; Costes, Laurence; Coulter, Tanya; Courteille, Virginie; Cristina, Maria; Cucuruz, Maria; Dabrowska, Leonik Nel; Dabrowska Leonik, Nel; Dähling, Mandy; Daly, Mary Louise; Daniel, Claudia-Sabrina; Danieli, Maria Giovanna; Darroch, James; Davies, Graham; De Gracia Roldan, Javier; de Nadai, Narimene; de Schutter, Iris; De Vergnes, Nathalie; de Vries, Esther; de Witte, Josine; deBaets, Frans; Debert, Theo; DeBoeck, Christiane; Defila, Corina; Deimel, Judith; Delaplace, Diane; Dellepiane, Rosa Maria; Dellert, Nelli; Delliera, Laura; Delor, Anita; Demel, Ulrike; Dempster, John; den Os, M.M.; Dengg, Rosmarie; Desisa, Sora Asfaw; Desta, Alexandra; Detkova, Drahomira; Dewerchin, Maite; Dieli Crimi, Romina; Dilloo, Dagmar; Dimitriou, Florentia; Dinges, Sarah Svenja; Dinser, Jasmin; Dipani, Nabila; Dirks, Johannes; Dittrich, Anna-Maria; Djermane, Lylia; Dogru, Yagmur; Dogu Esin, Figen; Dombrowski, Angelika; Dominguez Escobar, Julia; Döring, Michaela; Drabe, Camilla Heldbjerg; Drerup, Susann; Drexel, Barbara; Driessen, G.J.A.; Dückers, Gregor; Dudoit, Yasmine; Duppenthaler, Andrea; Ebetsberger-Dachs, Georg; Ecser, Mate Barnabas; Edgar, J. David; Eekman, Maartje; Ehl, Stephan; Ehrat, Rosanna; Eisl, Eva; Ekwall, Olov; El Hawary, Rabab; El-Helou, Sabine M.; El-Marsafy, Aisha; Elbe, Sarina; Elcombe, Suzanne; Eldash, Alia; Eldeen, Youssif Alkady Radwa Salah; Ellerbroek, P.M.; Elling, Roland; Elliott, Jane; Elmarsafy, Aisha; Engelhardt, Angelika; Ernst, Diana; Ersoy, Fügen; Esper, Stefanie; Esteves, Isabel; Etzioni, Amos; Exley, Andrew; Faber, Martin; Fabio, Giovanna; Fahrni, Gaby; Faletti, Laura Eva; Faletti, Laura Eva; Farber, Claire-Michele; Farela Neves, João; Faria, Emilia; Farkas, Henriette; Farmaki, Evangelia; Faßhauer, Maria; Fasth, Anders; Fätkenheuer, Gerd; Faustmann, Stefanie; Fecker, Gisela; Feighery, Conleth; Feiterna-Sperling, Cornelia; Fernandez-Cruz, Perez Eduardo; Ferreira Concalo, Cordeiro; Ferster, Alice; Feuchtinger, Tobias; Feyen, Oliver; Finke, Daniela; Fischer, Alain; Fitter, Sigrid; Flaschberger, Stefan; Fleckenstein, Lucia; Föll, Dirk; Fontana, Adriano; Forino, Concetta; Förster-Waldl, Elisabeth; Franchet, Nora; Freitag, Dagmar; Frey, Urs P.; Frick, Hannah Margarete; Friedel, Elisabeth; Friedrich, Wilhelm; Frisch, Barbara; Frischknecht, Lukas; Fritzemeyer, Stephanie; Gagro, Alenka; Gahr, Manfred; Galal, Nermeen Mouftah; Gambineri, Eleonora; Gamper, Agnes; Gams, Franziska; Ganzow, Astrid; Garcelon, Nicolas; Garcez, Tomaz; GarciaPrat, Marina; Gardulf, Ann; Garibay, Janine; Garwer, Birgit; Gathmann, Jonathan; Gathmann, Benjamin; Gebauer, Corinna; Geberzahn, Linda; Geikowski, Tilman; Geisen, Ulf; Gemander, Christiane; Gennery, Andrew R.; Gerisch, Marie; Gernert, Michael; Gerrer, Katrin; Gerschmann, Stev; Ghosh, Sujal; Giannini, Carolin; Gil Herrera, Juana; Gimenez Sanz, Noemi; Girndt, Matthias; Girrbach, Ramona; Girschick, Hermann; Gkantaras, Antonios; Gkougkourelas, Ioannis; Gładysz, Dominika; Gnatowski, Susanne; Gnodtke, Elisabeth; Goda, Vera; Goddard, Sarah; Goebel, Daniela; Goffard, Jean-Christophe; Goldacker, Sigune; Gollowitsch, Eva Maria; Gomes, Manuella; Gompels, Mark; González, Míriam; Gonzalez Granado, Luis Ignacio; Gordins, Pavels; Gößling, Katharina; Göschl, Lisa; Gossens, Lucy; Gowin, Ewelina; Graafen, Lea; Graca, Leo; Gradauskiene-Sitkauskiene, Brigita; Graf, Dagmar; Graf, Norbert; Grange, Elliot; Grashoff, H.Anne; Greil, Johann; Grigoriadou, Sofia; Grimbacher, Bodo; Gronlund, Helen; Groß-Wieltsch, Ute; Guerra, Teresa; Guevara-Hoyer, Kissy; Gueye, Mor Seny; GueyeMor, Seny; Guibert, Noemie; Gülnur, Birgit; Güngör, Tayfun; Guseva, Marina; Guzman, David; Haag, Marcel; Haase, Gabriele; Haenicke, Henriette; Haerynck, Filomeen; Hafsa, Ines; Hagin, David; Haliti, Emine; Hallek, Michael; Hammarstroem, Lennart; Hancioglu, Gonca; Handgretinger, Rupert; Hanitsch, Leif G.; Hansen, Susanne; Hariyan, Tetyana; Harrer, Thomas; Hassunah, Pia; Hatzistilianou, Maria; Hauck, Fabian; Hauser, Thomas; Haverkamp, Margje H.; Hayman, Grant; Heath, Paul; Hedrich, Christian; Heeg, Maximilian; Heike, Michael; Heimbrodt, Martin; Heine, Sabine; Heininger, Ulrich; Heinrich, Christian; Heinz, Valerie; Heitger, Andreas; Helbert, Matthew; Helbling, Arthur; Heldbjerg Drabe, Camilla; Hellige, Antje; Hempel, Julya; Henderson, Karen; Henes, Jörg; Henneke, Philipp; Hennig, Christian; Henrichs, Karin; Herbst, Martin; Hermann, Walter; Hernandez, Manuel; Hernández, Anja; Heropolitanska-Pliszka, Edyta; Herriot, Richard; Herrmann, Friedrich; Herwadkar, Archana; Hess, Christoph; Hess, Ursula; Hesse, Sebastian; Higgins, Sonja; Hilfanova, Anna; Hilpert, Sophie; Hintze, Chantal; Hlaváčková, Eva; Hodl, Isabel; Hodzic, Adna; Hoernes, Miriam; Hoffmann, Christina; Holbro, Andreas; Höllinger, Christiane; Holtsch, Lisa; Holzer, Ursula; Holzinger, Dirk; Hönig, Manfred; Hönscheid, Andrea; Horn, Julia; Horneff, Gerd; Hoyoux, Claire; Hristova, Nataliya; Hübel, Kai; Hübner, Angela; Huemer, Christian; Huissoon, Aarnoud; Hülsmann, Brigitte; Hundsdörfer, Patrick; Huppertz, Hans-Iko; Huß, Kristina; Hussain, Sadia; Husson, Julien; Hüttner-Foehlisch, Tanja; Ijspeert, Hanna; Ikinciogullari, Aydan; ilknur, Kökçü; Irga, Ninela; Jablonka, Alexandra; Jahnz-Rozyk, Karina; Jakob, Marcus; Jakoby, Donate; Jakoby-Gaide, Donate; Jandus, Peter; Jansson, Annette; Jaquet, Melanie; Jardefors, Helene; Jargulinska, Edyta; Jauk, Barbara; Jesenak, Milos; Jilka, Katharina; Jolles, Stephen; Jones, Alison; Jones, Regina; Jonkman-Berk, Birgit; Jönsson, Göran; Joyce, Hilary J.; Juliana, Pricillia; Kabesch, Michael; Kager, Leo; Kahlert, Christian; Kaiser-Labusch, Petra; Kakkas, Ioannis; Kamitz, Dirk; Kanariou, Maria; Kanz, Lothar; Karakoc-Aydiner, Elif; Karanovic, Boris; Kartal-Kaess, Mutlu; Käser, Elisabeth; Katzenstein, Terese L.; Kayserova, Hana; Kelleher, Peter; Kerre, Tessa; Kilic, Sara Sebnem; Kindle, Gerhard; Kirchner, Martina; Kiwit, Simone; Kiykim, Ayca; Klasen, Jessica; Klaudel-Dreszler, Maja; Klein, Ariane; Klein, Christoph; Klein-Franke, Andreas; Kleine, Ilona; Kleinert, Stefan; Klemann, Christian; Klima, Marion; Klocperk, Adam; Kobbe, Robin; Kocacik Uygun, Dilara Fatma; Koch, Melanie; Kochler, Yvonne; Kohistani, Naschla; Kojic, Marina; Kolios, Antonio; Kölsch, Uwe; Koltan, Sylwia; Kondratenko, Irina; Königs, Christoph; Konoplyannikova, Julia; Kopac, Peter; Kopp, Jana; Körholz, Dieter; Körholz, Julia; Korte, Pauline; Kostyuchenko, Larysa; Kötter, Ina; Kracker, Sven; Králícková, Pavlina; Kramm, Christof; Kramme, Philipp; Krausz, Máté; Kreuz, Wolfhart; Krista, Johanna; Kriván, Gergely; Kropshofer, Gabriele; Krüger, Renate; Krystufkova, Olga; Ktistaki, Maria; Kühl, Jörn-Sven; Kühn, Alexander; Kuijpers, Taco W.; Kuis, Wietse; Kullmann, Silke; Kulozik, Andreas; Kumararatne, Dinakantha; Kümmler, Ria; Kündgen, Andrea; Kurenko-Deptuch, Magdalena; KussPaula, Cosima; Kütükcüler, Necil; Lafoix-Mignot, Cécile; Lamers, Beate; Lanbeck, Peter; Landais, Paul; Landwehr-Kenzel, Sybille; Langemeyer, Vanessa; Langer, Thorsten; Lankisch, Petra; Lanz, Nadia; Lara, Manrique de; Lara-Villacanas, Eusebia; Laubenthal, Lisa; Laws, Hans-Jürgen; Leahy, Ronan; Lee, Jae-Yun; Lehmann, Andrea; Lehmberg, Kai; Lehner, Patricia; Leibfrit, Hans; Leistner, Leoni; LeMignot, Loic; Lesch, Petra; Leutner, Simon; Liatsis, Manolis; Libai Véghová, Linda; Liebel, Johanna; Liese, Johannes G.; Linauskiene, Kotryna; Linde, Richard; Linßner, Martina; Lippert, Conrad Ferdinand; Litzman, Jiri; Llobet, Pilar; Lo, Babacar; Lodin, Tariq; Lokaj, Jindrich; Longhino, David; Longhurst, Hilary; Lopes da Silva, Susana; Lorenzen, Catharina; Lougaris, Vassilios; Löw, Doris; Lubatschofski, Annelie; Lucas, Mary; Lutz-Wiegers, Verena; Maaß, Sabine; Maccari, Maria Elena; Macura-Biegun, Anna; Maerz, Vanessa; Maggina, Paraskevi; Mahlaoui, Nizar; Mahrenholz, Hannah; Maier, Sarah; Makhlouf, Mounia; Malfroot, Anne; Malinauskiene, Laura; Mannhardt-Laakmann, Wilma; Manson, Ania; Mantkowski, Felicia; Manzey, Petra; Marasco, Carolina; Marcus-Mandelblit, Nufar; Marg, Wolfgang; Markelj, Gasper; Marodi, Laszlo; Marques, José Goncalo; Marques, Laura; Marschall, Karin; Martínez, Natalia; Martinez de la Ossa Saenz-Lopez, Rafael; Martinez-Saguer, Inmaculada; Martire, Baldassarre; Marzollo, Antonio; Masekela, Refiloe; Masjosthusmann, Katja; Masmas, Tania Nicole; Matamoros, Nuria; Mattern, Jutta; Mau-Asam, Pearl; McDermott, Elizabeth; McIntosh, Nichole; Meglic, Karmen Mesko; Meijer, Ruben; Meinhardt, Andrea; Meshaal, Safa; Messaoud, Yasmina; Meyer, Björn; Meyer-Olson, Dirk; Meyts, Isabelle; Micol, Romain; Micoloc, Bozena; Mielke, Gudrun; Milito, Cinzia; Milota, Tomas; Misbah, Siraj; Mödden, Carolin; Mohr, Michael; Mohrmann, Karina; Moin, Mostafa; Molinos, Luis; Möller, Jana; Möller-Nehring, Sarah; Morbach, Henner; Moschese, Viviana; Moser, Olga; Moshous, Despina; Motkowski, Radoslaw; Motwani, Jayashree; MouftahGalal, Nermeen; Müglich, Carmen; Mukhina, Anna; Muller, Eva; Müller, Christiane; Müller, Gabriele; Müller, Hedi; Müller, Ingo; Müller, Thomas; Müller, Zoe; Müller-Ladner, Ulf; Müller-Stöver, Sarah; Münstermann, Esther; Murtra Garrell, Núria; Muschaweck, Moritz; Mutert, Miriam; Nademi, Zohreh; Naik, Paru; Näke, Andrea; Nalda, Andrea Martin; Nasrullayeva, Gulnara; Naumann-Bartsch, Nora; Nemitz, Verena; Neth, Olaf; Neubauer, Andreas; Neubert, Jennifer; Neumann, Carla; Niehues, Tim; Niemuth, Mara; Nieters, Alexandra; Nieuwhof, Chris; Nolkemper, Daniela; Noorani, Sadia; Noorlander, Budde Adya; Notarangelo, Luigi D; Notheis, Gundula; Nowatsh, Sanam Amelie; Obenga, Gaelle; Ocak, Suheyla; Oker, Mehmet; Olbrich, Peter; Olipra, Anna; Omran, Heymut; Oommen, Prasad; Opitz, Linda; Orosova, Jaroslava; Oskarsdottir, Solveig; Özsahin, Hülya; Pac, Malgorzata; Pachlopnik-Schmid, Jana; Pandolfi, Franco; Papadopoulou-Alataki, Efimia; Papastamatiou, Theodora; Papatriantafillou-Schmieder, Anna; Parra-Martinez, Alba; Paschenko, Olga; Pašić, Srdjan; Pasnik, Jarek; Patel, Smita; Pavlík, Martin; Paz Artal, Estela; Peeters, Anouk; Pereira da Silva, Sara Branco; Perez-Becker, Ruy; Perez-Guzman, Marc; Pergent, Martine; Perlhagen, Markus; Peter, Hans-Hartmut; Petrić, Marin; Pfreundschuh, Michael; Philippet, Pierre; Picard, Capucine; Pietrucha, Barbara; Pietzsch, Leonora; Pignata, Claudio; Piquer Gibert, Monica; Pirolt, Kerstin; Plebani, Alessandro; Pleguezuelo, Daniel E.; Pollok, Katrin; Pommerening, Helena; Popihn, Daniela; Poplonek, Aleksandra; Popp, Marina; Porta, Fulvio; Portegys, Jan; Posfay-Barbe, Klara; Potjewijd, Judith; Poulheim, Sebastian; Prader, Seraina; Prämassing-Scherzer, Petra; Prelog, Martina; Prevot, Johan; Price, Arthur; Price, Timothy; Proesmans, Marijke; Provot, Johan; Pulvirenti, Federica; Quinti, Isabella; Raab, Anna; Rack, Anita; Raffac, Stefan; Ramos Oviedo, Eduardo; Randrianomenjanahary, Philippe; Ranohavimparany, Anja; Raptaki, Maria; Rashidzadeh, Roonaka; Rathwallner, Margit; Reda, Shereen; Redouane, Nahida; Regateiro, Frederico S.; Reichenbach, Janine; Reimers, Bianca; Reinhardt, Cornelia; Reinhardt, Dirk; Reinprecht, Anne; Reiß, Tamara; Reisli, Ismail; Renner, Eleonore; Rezaei, Nima; Richter, Alex; Richter, Darko; Rieber, Nikolaus Peter; Rieckehr, Nadja; Riedel, Marion; Riescher, Heidi; Rischewski, Johannes; Ristl, Nicole; Ritterbusch, Henrike; Ritz, Tanja; Rivier, Francois; Robinson, Peter; Rockstroh, Jürgen K.; Roesler, Joachim; Rofiah, Himatur; Rogerson, Elizabeth; Rolfes, Elisabeth; Roller, Beate; Romanyshyn, Yaryna; Rondelli, Roberto; Roosens, Fien; Rösen-Wolff, Angela; Rösler, Valentina; Roth, Johannes; Rothoeft, Tobias; Roubertie, Agathe; Rübsam, Gesa; Rusch, Stephan; Rutgers, Abraham; Ryan, Paul; Sach, Gudrun; Sadeghi, Kambis; Sahrbacher, Ulrike; Saidi, Angelika; Sanal Tezcan, Özden; Sanchez-Ramon, Silvia; Santos, Juan Luis; Sargur, Ravishankar; Savchak, Ihor; Savic, Sinisa; Schaaf, Bernhard; Schaefer, Marzena; Schäfe, Christina; Scharbatke, Eva; Schatorje, Ellen; Schauer, Uwe; Scheibenbogen, Carmen; Scheible, Raphael; Scheinecker, Clemens; Schiller, Romana; Schilling, Beatrice; Schilling, Freimut; Schlieben, Steffi; Schmalbach, Thilo; Schmalzing, Marc Thomas; Schmid, Pirmin; Schmidt, Nadine; Schmidt, Reinhold Ernst; Schmitz, Monika; Schneider, Dominik; Schneider, Dominik T.; Schneppenheim, Reinhard; Scholtes, Cathy; Schölvinck, E.H.; Schönberger, Stefan; Schreiber, Stefan; Schrijvers, Rik; Schroll, Andrea; Schruhl, Simone; Schrum, Johanna; Schubert, Ralf; Schuetz, Catharina; Schuh, Sebastian; Schulz, Ansgar; Schulz, Claudia; Schulze, Ilka; Schulze-Koops, Hendrik; Schulze-Sturm, Ulf; Schumacher, Eva-Maria; Schürmann, Elvira; Schürmann, Gesine; Schuster, Volker; Schwaneck, Eva; Schwarz, Klaus; Schwarz, Tobias; Schwarze-Zander, Carolynne; Schweigerer, Lothar; Sediva, Anna; Seebach, Jörg; Seger, Reinhard; Segerer, Florian; Seidel, Markus G.; Selle, Barbara; Seneviratne, Suranjith; Seppänen, Mikko; Shabanaj, Hatidje; Sharapova, Svetlana; Shcherbina, Anna; Shillitoe, Benjamin; Siepelmeyer, Anne; Siepermann, Kathrin; Simon, Anna; Simon, Arne; Simon-Klingenstein, Katja; Simonovic, Marija; Simsen-Baratault, Merlin; Sindram, Elena; Skapenko, Alla; Skarke, Maiken; Skomska-Pawliszak, Malgorzata; Slatter, Mary; Smet, Julie; Smith, C. I. Edvard; Sobh, Ali; Sobik, Bettina; Sogkas, Georgios; Sohm, Michael; Solanich, Xavier; Solanich-Moreno, Xavier; Soler Palacín, Pere; Sollinger, Franz; Somech, Raz; Sonnenschein, Anja; Soresina, Annarosa; Sornsakrin, Marijke; Soura, Stavrieta; Spaccarotella, Sabrina; Spadaro, Guiseppe; Sparber-Sauer, Monika; Specker, Christof; Speckmann, Carsten; Speidel, Lisa; Speletas, Matthaios; Stachel, Klaus-Daniel; Stadon, Catherine; Stanislas, Aurélie; Stapornwongkul, Cynthia; Staus, Paulina; Steck, Regina; Steele, Cathal; Steffin, Herbert; Steiner, Urs; Steinmann, Sandra; Stevens, Wim; Stiefel, Martina; Stieger, Sarah; Stiehler, Sophie; Stimm, Hermann; Stojanov, Silvia; Stoll, Matthias; Stoppa-Lyonnet, Dominique; Strapatsas, Tobias; Strauß, Gabriele; Streiter, Monika; Strik-Albers, Riet; Strotmann, Gaby; SuárezCasado, Héctor; Subiza, Jose Luis; Sundin, Mikael; Süß, Birgit; Sutter, Fabienne; Szaflarska, Anna; Szemkus, Monika; Tamary, Hannah; Tantou, Sofia; Tarzi, Michael D.; Taschner, Helga; Tedgard, Ulf; Teixeira, Carla; ten Berge, RJM; Tenbrock, Klaus; Tester, Sabine; Tezcan, Ilhan; Thalguter, Sonja; Thalhammer, Julian; Thoma, Katharina; Thomas, Moira; Thomczyk, Fabian; Thon, Vojtech; Thrasher, Adrian; Tierney, Patricia; Tietsch, Nadine; Tommasini, Alberto; Tönnes, Beate; Tony, Hans-Peter; Trachana, Maria; Trapp, Carmen; Tricas, Lourdes; Trindade Neves, Maria Conceicao; Trischler, Jordis; Ubieto, Hugo; Uelzen, Anett; Uhlmann, Annette; Ullrich, Jan; Ullrich, Kurt; Ünal, Ekrem; Urbanski, Gerhard; Urschel, Simon; Uszynska, Aleksandra; Vacca, Angelo; Vaganov, N.N.; Vagedes, Daniel; Valicevic, Stefanie; Vallelian, Florence; van Beem, Rachel T; van Damme, Charlotte; van de Ven, Annick; van den Berg, J. Merlijn; van der Flier, Michiel; van Dissel, J.T.; van Hagen, P.M.; van Montfrans, J.M.; van Ogtrop, Geoffrey; van Rens, Jacqui; van Riel, Christel A.M.P.; van Royen-Kerkhof, Annet; van Well, G.Th.J.; Vasiliki, Antari; Velbri, Sirje; Vencken, Jo; Ventura, Alessandro; Vermeulen, François; Vermylen, Christiane; Viemann, Dorothee; Viereck, Anja; Villa, Anna; Vincke, Jeroen; Vinnemeier-Laubenthal, Lisa; Vo Thi, Kim Duy; Voeller, Mirjam; Vollbach, Kristina; Volokha, Alla; von Bernuth, Horst; von Bismarck, Philipp; Voss, Rebecca; Voß, Sandra; Vural, Yüksel; Wachuga, Heike; Wagner, Norbert; Wagström, Per; Wahle, Matthias; Wahn, Volker; Wapp, Nadine; Warnatz, Klaus; Warneke, Monika; Warris, Adilia; Wasmuth, Jan-Christian; Wasserfallen, Jean-Blaise; Wawer, Angela; Weber, Manfred; Weemaes, Corrie MR; Wege, Lisa; Wehrle, Julius; Weidinger, Stephan; Weiß, Michael; Weißbarth-Riedel, Elisabeth; Werner, Antje; Wessel, Sara; Westkemper, Marco; Wicher, Monika; Wickmann, Lutz; Wiegert, Sabine; Wiehe, Monique; Wiehler, Katharina; Wiesböck, Lydia; Wiesik-Szewczyk, Ewa; Williams, Anthony; Winkler, Beate; Winkler, Christel; Winkler, Martina; Winkler, Melanie; Wintergerst, Uwe; Wisgrill, Lukas; Witte, Torsten; Wittkowski, Helmut; Wolf, Barbara; Wölke, Sandra; Wolschner, Christina; Wolska-Kusnierz, Beata; Wood, Philip; Workman, Sarita; Worth, Austen; Wortmann, Michaela; Wuillemin, Walter Alfred; Wulffraat, Nico M; Wustrau, Katharina; Wyndham-Thomas, Chloé; Yegin, Olcay; Yildiran, Alisan; Yilmaz, Denise; Young, Patrick; Yucel, Esra; Zečević, Milica; Želimir, Erić; Zepp, Fred; Zetzsche, Klaus; Zeuner, Rainald; Zielen, Stefan; Zimmermann, Martina; **AND additional malignancy management L2 survey collaborators**: Baselli, Lucia; Bobcakova, Anna; Deripapa, Elena; Gabzdilova, Juliana; Gadziev, Alibey; Heldbjerg-Drabe, Camilla; Kuehnle, Ingrid; Lodi, Lorenzo; Markocsy, Adam; Martin, Andrea; Petrovic, Otilia; Ricci, Silvia; Rivalta, Beatrice; Rodina, Yulia; Sarli, Walter Maria; Strauss, Timmy.

## ONLINE REPOSITORY

## SUPPLEMENTARY METHODS

### Level 2 survey questions

A questionnaire was used to obtain additional information regarding malignancies in IEI patients within the ESID registry. The following questions and answer options (if applicable) were presented to the respondents.

#### General

- ESID registry ID of the patient. (free text)

#### Malignancy diagnosis

- Date of diagnosis of the malignancy? (dd/mm/yyyy)
- Age (in years) at diagnosis of the malignancy? (free text)
- Please indicate the type of malignancy.
  - Leukemia
  - Lymphoma
  - Solid tumor (incl. CNS tumors)*
  - Other

*CNS: central nervous system

- In case of leukemia, what was the risk group at diagnosis?
  - Standard risk
  - Intermediate risk
  - High risk
  - Unknown
- In case of lymphoma, what was the stage of disease at diagnosis?
  - Stage I
  - Stage II
  - Stage III
  - Stage IV
  - Unknown
- In case of a solid tumor (incl. CNS tumors), what was the stage of disease at diagnosis?
  - Localized
  - Metastatic
  - Unknown
- What “other” kind of malignancy was diagnosed? (free text)
- What was the cytogenetics or molecular genetics of the malignancy at diagnosis (if determined). (free text)

#### Other (non-IEI) risk factors for developing malignancy

- How likely do you consider the possibility that lifestyle factors (e.g. tabacco, alcohol, sun exposure, other) contributed to the cancer development in this patient?
  - Very likely
  - Likely
  - Unlikely
  - Excluded
- How likely do you consider the possibility that the malignancy is associated to previous (long-term) treatment with immune suppressive or immune modulating drugs for the IEI?
  - Very likely
  - Likely
  - Unlikely
  - Excluded
- How likely do you consider the possibility that the development of the malignancy is linked to chronic infection with a documented oncogenic/transforming pathogen (e.g. EBV, HCV, HPV, or Helicobacter) in this patient?
  - Very likely
  - Likely
  - Unlikely
  - Excluded
- How likely do you consider the possibility that the malignancy is associated with chronic inflammation in this patient?
  - Very likely
  - Likely
  - Unlikely
  - Excluded
- Was the malignancy diagnosed after the patient had undergone HSCT?
  - Yes
  - No
  - Unknown
- Did the second/third malignancy develop in a prior radiotherapy field?*
  - Yes
  - No
  - Unknown

*This question was only given in case of a second or third primary malignancy; see below.

- Is there a positive family history of cancer independent of a confirmed IEI?
  - Yes
  - No
  - Unknown
- In case of a positive family history of cancer, what degree of relatives developed cancer?
  - 1^st^ degree
  - 2^nd^ degree
  - 3^rd^ degree
  - 4^th^ or higher degree
  - Unknown

#### Treatment

- What kind of oncological treatment did the patient receive?
  - Standard treatment protocol according to the type of malignancy, without adaptations
  - Standard, with dose modifications during therapy based on encountered toxicities (i.e. secondary quantitative modifications)
  - Standard, with upfront quantitative modifications out of caution (i.e. primary quantitative modifications)
  - Individualized treatment based on the IEI (i.e. primary quantitative or qualitative modifications, e.g. dose reductions, no radiotherapy, no alkylating agents)
  - Individualized treatment for reasons other than the IEI (i.e. primary quantitative or qualitative modifications, e.g. dose reductions, no radiotherapy)
  - Unknown
- In case of treatment modifications/individualized treatment: how were these modifications decided upon?
  - Local oncological team decision
  - Local multidisciplinary team decision (incl. oncologists and immunologists)
  - National or international oncological expert opinion
  - National or international multidisciplinary expert opinion
  - Unknown
- In case of treatment modifications/individualized treatment, was the decision based on published literature/guidelines specific for this IEI and malignancy?
  - Yes
  - No
  - Unknown
- Was an immunologist involved during the time of the cancer therapy?
  - Yes
  - No
  - Not applicable, immunologist and oncologist are the same physician
  - Unknown
- Did the immunological treatment change during the time of the oncological treatment?
  - Yes
  - No
  - Unknown
- Did the patient receive radiotherapy as part of the oncological treatment?
  - Yes, full dose radiotherapy according to standard of care for this type of malignancy
  - Yes, full dose but radiotherapy is not standard of care for this type of malignancy
  - Yes, but the radiotherapy dose was reduced in this patient because of the risk of excessive toxicity
  - No, because radiotherapy is not standard of care for this type of malignancy
  - No, radiotherapy was not done in this patient because of the risk of excessive toxicity
  - Unknown
- Did the patient receive immune therapy as part of the oncological treatment?
  - Yes
  - No
  - Unknown
- Did the underlying IEI influence the decision to give or omit immune therapy?
  - No
  - Yes, the IEI was an argument pro immune therapy
  - Yes, the IEI was an argument against immune therapy
  - Unknown
- Was allogeneic HSCT performed after or as part of the oncological treatment?
  - Yes
  - No
  - Unknown
- Did the underlying IEI influence the decision to perform or not to perform HSCT?
  - No
  - Yes, the IEI was an argument pro HSCT
  - Yes, the IEI was an argument against HSCT
  - Unknown
- Did the underlying IEI have an indication for HSCT?
  - Yes
  - No
  - Unknown
- In case HSCT was performed, what was the malignancy remission status prior to HSCT?
  - HSCT in CR1
  - HSCT in CR2 or higher
  - No remission
  - Unknown
- In case HSCT was performed, what type of conditioning regimen was used?
  - Myeloablative conditioning
  - Non-myeloablative conditioning
  - Unknown

#### Complications

- Do you think the underlying IEI prolonged the regular treatment period for the malignancy?
  - Yes
  - No
  - Unknown
- What type/intensity of antimicrobial prophylaxis (antibiotics, antifungals, antivirals, immunoglobulins) was used during the oncological treatment?
  - According to standard local guidelines
  - Intensified prophylaxis based on the underlying IEI
  - Intensified prophylaxis for reasons other than the underlying IEI
  - Unknown
- Do you think the patient experienced more severe complications of any kind (e.g., requiring ICU admission, organ failure, etc.) in comparison to cancer patients without underlying IEI?
  - Yes
  - No
  - Unknown
- Do you estimate that the patient had more frequent and/or more severe infections during the oncological treatment in comparison to cancer patients without underlying IEI?
  - Yes
  - No
  - Unknown
- Did the patient develop a life-threatening infectious complication during the oncological treatment requiring intensive care (ICU)?
  - Yes
  - No
  - Unknown
- Did the patient develop a grade 3-4 inflammatory/autoimmune complication during the oncological treatment (including unexplained systemic inflammation)?
  - Yes
  - No
  - Unknown
- Did the patient develop any other grade 3-4 toxicity (non-infectious, non-inflammatory/autoimmune) during the oncological treatment?
  - Yes
  - No
  - Unknown
- In case of another grade 3-4 toxicity (non-infectious, non-inflammatory/autoimmune), please specify the organ/system. (free text)

#### Outcome

- What is the current survival status of the patient?
  - Alive
  - Deceased
  - Unknown
- Age (in years) of last follow-up? (free text)
- Date of last follow-up? (dd/mm/yyyy)
- What is the current age (in years) of the patient (if alive)? (free text)
- What is the current disease state regarding the malignancy?
  - In remission
  - Stable disease
  - Refractory disease
  - Relapse
  - Death of disease (1^st^ malignancy)*
  - Death due to second/third malignancy*

* These answer options were only given in case of a second or third primary malignancy; see below.

- In case of relapse of the malignancy: what was the timing of relapse?
  - No remission achieved, refractory disease
  - Very early relapse, i.e. < 18 months after diagnosis and/or < 6 months after completion of primary therapy
  - Early relapse, i.e. between 18 and 30 months after diagnosis
  - Late relapse, i.e. ≥ 30 months after diagnosis and ≥ 6 months after completion of primary therapy
- In case the patient has died, what was the age (in years) at time of death? (free text)
- In case the patient has died, what was the cause of death?
  - Death related to (progression of) the malignancy
  - Death related to toxicity of the oncological treatment
  - Death related to an IEI-associated complication
  - Death caused by another malignancy
  - Other cause of death
- Do you assume the IEI adversely affected the outcome of the malignancy?
  - Yes
  - No
  - Unknown
- If you assume the IEI adversely affected the outcome of the malignancy, what do you think was the reason for this? (multiple answers could be selected)
  - Related to the underlying IEI (infections)
  - Related to the underlying IEI (immune dysregulation)
  - Related to pretreatment and comorbidity
  - The maximum oncological therapy could not be given
  - Other

#### Follow-up

- After successful treatment of the malignancy, where is the follow-up performed?
  - At the oncological clinic (including late effects/survivorship clinic)
  - At the immunology clinic
  - At a combination of the oncological and immunological clinic
  - Unknown
  - Not applicable (no remission achieved)
- After successful treatment of the malignancy, was the prophylactic oncological screening intensified in this patient?
  - Yes
  - No
  - Unknown
- If the prophylactic oncological screening post-malignancy was intensified in this patient, how was this done?
  - Increased frequency of surveillance examinations
  - Additional imaging/lab examinations
  - Both increased frequency of examinations and additional examinations
  - Other
  - Unknown
- Is there a prophylactic oncological screening guideline available at your clinic for this IEI in the POST-malignancy setting (in-house expert opinion, or guideline-based)?
  - Yes
  - No
  - Unknown
- Is there a prophylactic oncological screening guideline available at your clinic for this IEI in the PRE-malignancy setting (in-house expert opinion, or guideline-based)?
  - Yes
  - No
  - Unknown

#### General

- Did the patient develop another primary malignancy after this one (not relapse of previous primary malignancy, not secondary to previous treatment)?*
  - Yes
  - No
  - Unknown

* In case the patient had developed multiple primary malignancies, the respondents were asked to answer all questions for each primary malignancy separately (up to 3 malignancies per patient). The set of questions for the other primary malignancies appeared after completing the first one.

- How many primary malignancies (except relapses) did the patient suffer from in total?
  - One
  - Two
  - Three
  - More than three
- If you would like to provide any other relevant information on this patient in the context of the malignancy missed by the questionnaire, please do so here. (free text)
- Please score your satisfaction with this survey (1-100). (free text)
- Please repeat the ESID registry ID of the patient (for confirmation purposes). (free text)

## SUPPLEMENTARY RESULTS

### Malignancy risk factors non-intrinsic to the IEI reported in the survey cohort

The survey explored which factors non-intrinsic to the IEI may have contributed to the overall cancer risk (Supplementary Figure E3). In 11.8% of malignancies, lifestyle-related factors (e.g. tobacco, alcohol, sun exposure) were believed to have contributed to their development. In 9.9% of patients, a positive family history of cancer (independent of the IEI) was reported. Of these, respondents indicated that 47.4% had a first-degree and 39.5% a second-degree relative with cancer. Several factors indirectly linked to the IEI were also deemed to have influenced the cancer risk. 7.3% of malignancies were thought to be associated with previous treatment with immunosuppressive or immunomodulating drugs for the IEI. Chronic infection with a documented oncogenic or transforming pathogen was believed to contribute to cancer development in 26.4% of malignancies, whereas chronic inflammation was reported to have played a role in 30.7%. Furthermore, 3.9% of malignancies was reported to have developed after the patient had undergone a hematopoietic stem cell transplantation (HSCT). Finally, 12.2% of second or higher primary malignancies were reported to have developed in a prior radiotherapy field. Overall, in 24.1% of malignancies reported in the survey cohort, a combination of two or more of the above-mentioned factors were considered to have contributed to the cancer risk. In contrast, in 41.3% of malignancies, no additional risk factors – besides those intrinsic to the IEI – were believed to have played a role.

### Reported information on radiotherapy and immune therapy as part of the oncological treatment, and on antimicrobial prophylaxis and immunological therapy during oncological treatment in the survey cohort

A full dose of radiotherapy, as part of the oncological treatment, was reported to be given in 17.7% of malignancies. In contrast, in 7.8% of malignancies, radiotherapy was believed to be reduced or completely omitted because of risk of excessive toxicity. In 66.2% of malignancies, radiotherapy was not considered to be standard of care and therefore not part of the oncological treatment. Immune therapy was part of the oncological treatment in 14.4% of malignancies. In a minority of cases, the IEI was considered to have influenced the decision whether to give immune therapy, being an argument in favor of immune therapy in 5.6% and against it in 0.7%. According to the survey respondents, antimicrobial prophylaxis during oncological treatment was most often done according to standard local practices (51.3%) but intensified because of the IEI in 18.3%. In 30.4% of cases, the antimicrobial prophylaxis regimen was unknown. Immunological therapy was believed to be adapted in 15.6% of malignancy treatments, whereas it remained the same in 76.6%. In 7.8%, it was unknown whether changes to the immunological treatment had been made.

## SUPPLEMENTARY TABLES

**Table E1.** Distribution of malignancies (n=436) across the IEI categories reported in the survey cohort.

| IEI diagnosis<sup>o</sup> | Leukemia | Lymphoma | Solid tumor (incl. CNS) | Other (LCH) |
| --- | --- | --- | --- | --- |
|  | (number, percentage within IEI category) |  |  |  |
| Immunodeficiencies affecting cellular and humoral immunity | 2 (5.1%) | 22 (56.4%) | 14 (35.9%) | 1 (2.6%) |
| Combined immunodeficiencies with associated or syndromic features | 19 (18.4%) | 60 (58.3%) | 24 (23.3%) | 0 |
| Predominantly antibody deficiencies | 6 (2.7%) | 89 (40.5%) | 125 (56.8%) | 0 |
| Diseases of immune dysregulation | 2 (5.4%) | 26 (70.3%) | 9 (24.3%) | 0 |
| Congenital defects of phagocyte number or function | 2 (100%) | 0 | 0 | 0 |
| Defects in intrinsic and innate immunity | 0 | 1 (50%) | 1 (50%) | 0 |
| Autoinflammatory disorders | 1 (16.7%) | 1 (16.7%) | 4 (66.7%) | 0 |
| Complement deficiencies | 0 | 2 (33.3%) | 4 (66.7%) | 0 |
| Phenocopies of inborn errors of immunity | 1 (11.1%) | 1 (11.1%) | 7 (77.8%) | 0 |
| Unspecified | 4 (44.4%) | 5 (55.6%) | 0 | 0 |

<sup>o</sup>According to the IUIS classification<sup>1</sup>. IEI diagnosis was missing for two patients of whom one had two malignancies. CNS, central nervous system; IEI, inborn errors of immunity; LCH, Langerhans cell histiocytosis.

## SUPPLEMENTARY LEGENDS

**Figure E1.**
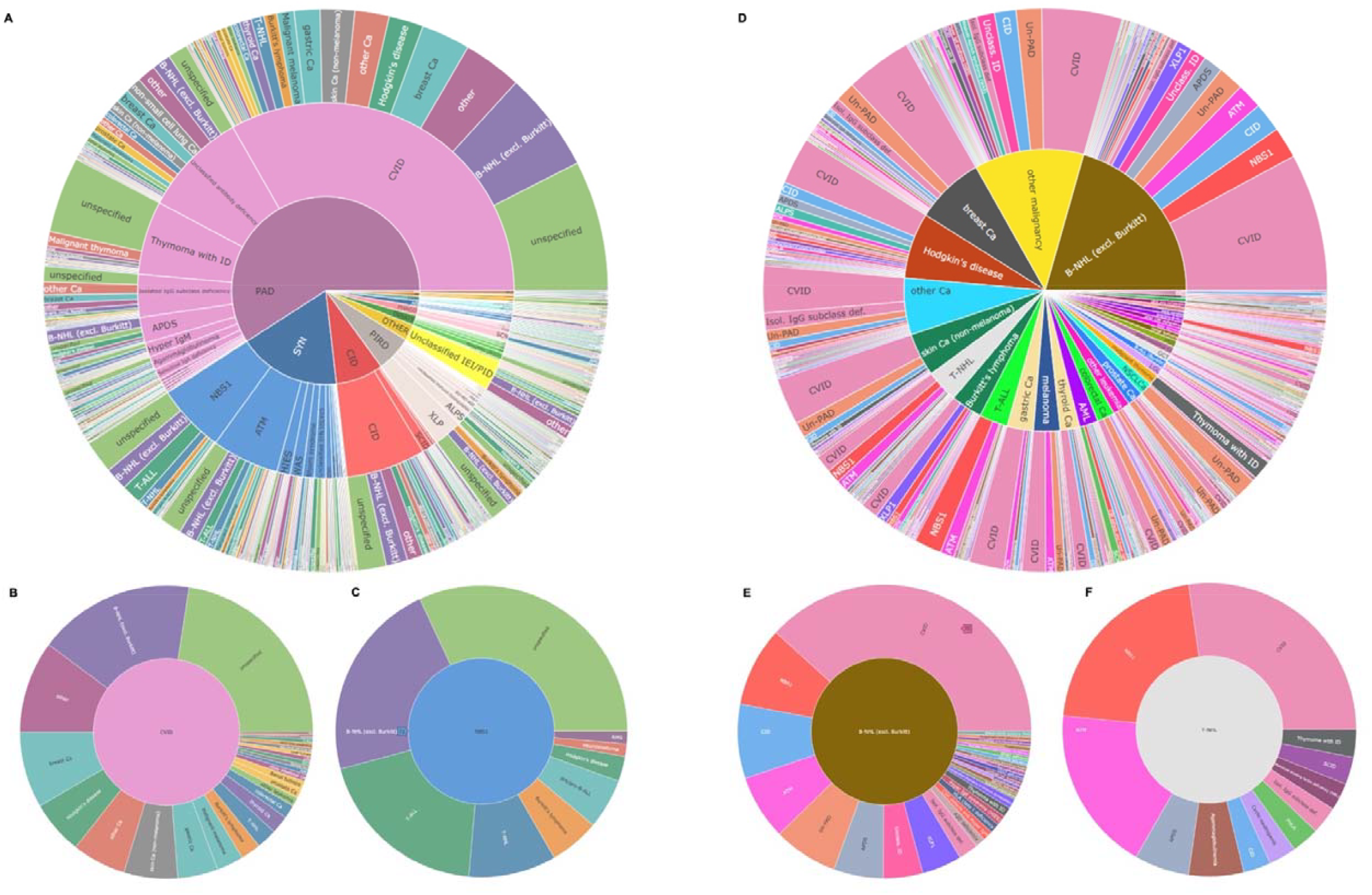
Hierarchical pie charts link detected malignancies with IEI/PID diagnoses of ESID level 1 registry patients. Screenshots of two online interactive figures showing this association (https://esid.org/html-pages/Suppl_figure_ESID_sunburst_malignancy_17.html AND https://esid.org/html-pages/Suppl_figure_ESID_sunburst_malignancy_rev_29.html) show, ***A***, the distribution of IEI/PID with reported malignancies according to the IUIS category of IEI/PID, diagnosis, and malignancy subtype, with an enlargement for CVID in ***B***, and of NBS in ***C***. The right panel, ***D***, shows the malignancy-centered view with an enlargement of B-NHL in ***E***, and of T-NHL with associated IEI/PID in ***F***. Data and numbers are the same as in Figure 1. Mouse-over in the interactive figure shows absolute numbers.

**Figure E2.**
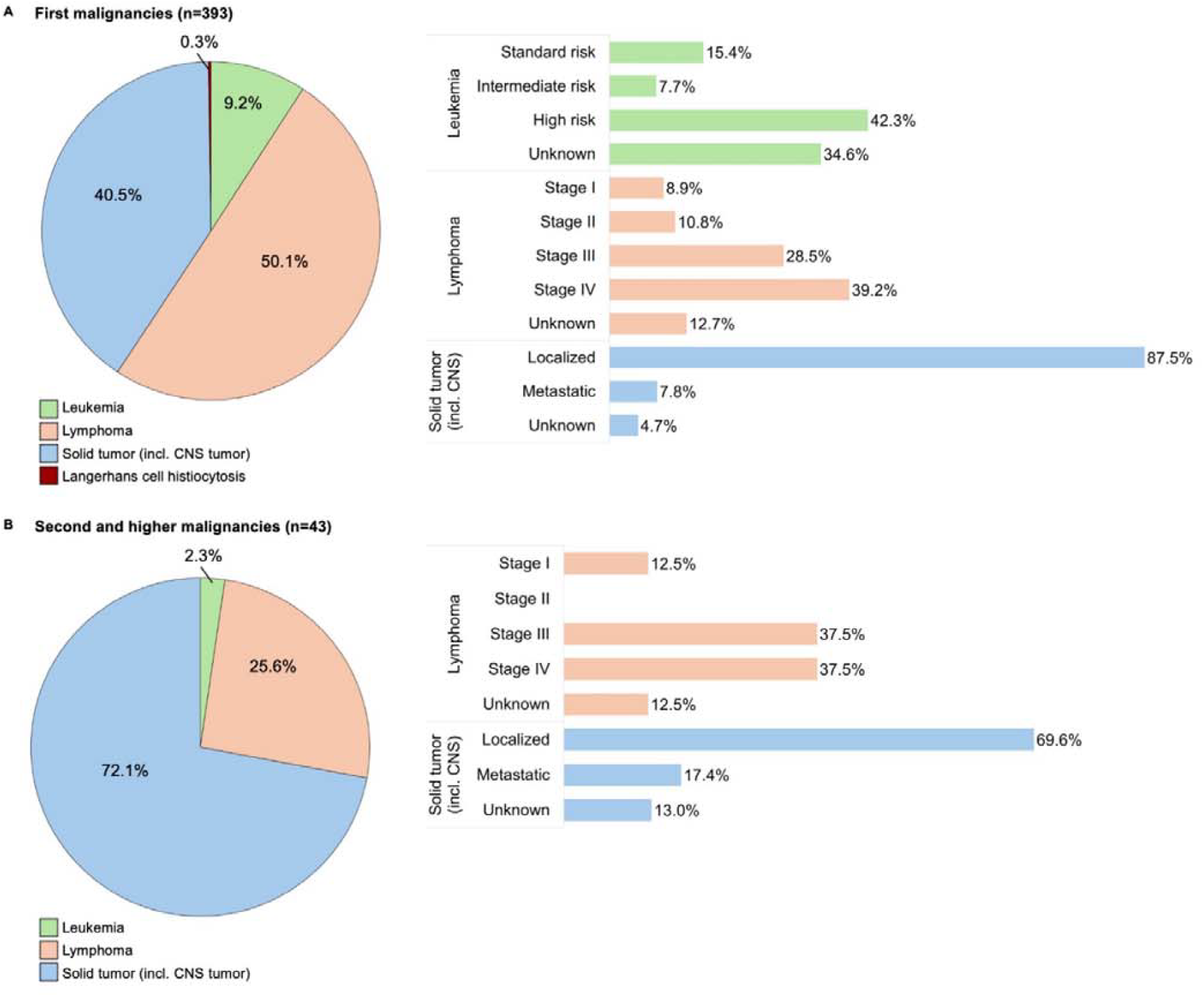
Overview of first malignancies (A) and second and higher malignancies (B) reported in the ESID registry level 2 sub-study (L2 survey) cohort. The pie charts depict the types of malignancies. The bar charts show the risk groups or disease stages of the malignancies at diagnosis. Note that there was only 1 leukemia reported as second or higher malignancy, for which the risk group at diagnosis was missing from the survey. CNS; central nervous system.

**Figure E3.**
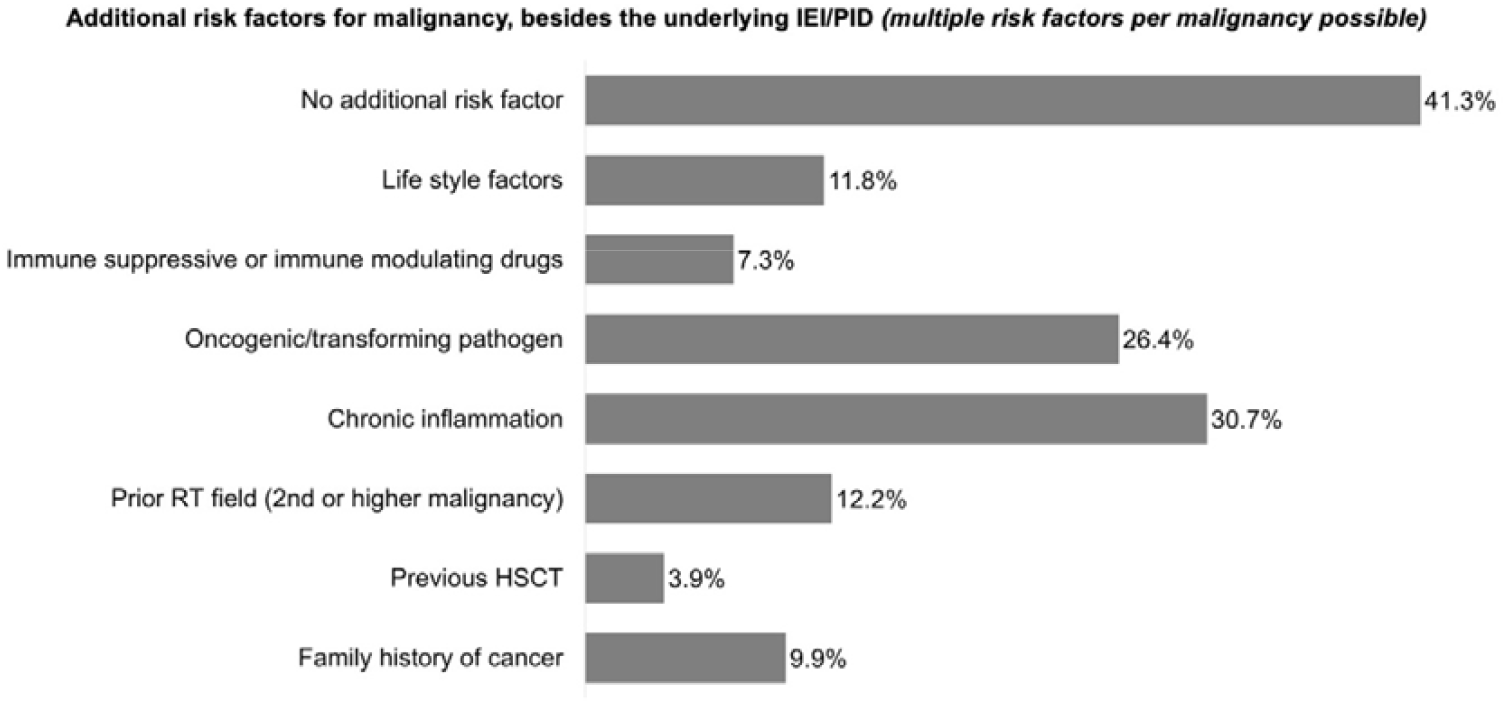
Malignancy risk factors, besides the underlying IEI/PID, reported in the L2 survey cohort. HSCT, hematopoietic stem cell transplantation; IEI, inborn errors of immunity; PID, primary immunodeficiency; RT, radiotherapy.

**Figure E4.**
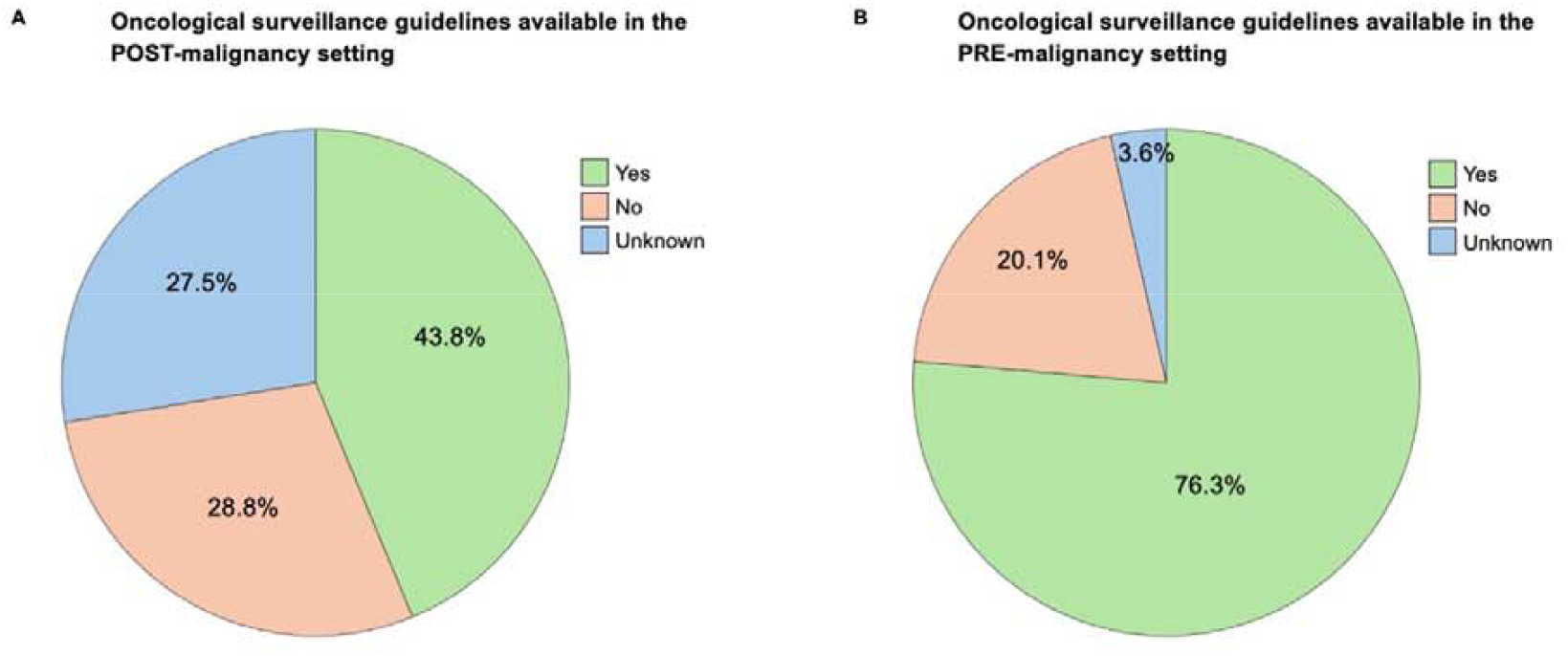
The availability of oncological surveillance guidelines specific for the IEI/PID subtype, as reported in the survey. A. In the setting in which a malignancy has already occurred. B. In the setting were the patient has not (yet) developed a malignancy.

